# Clot Twist – D-dimer analysis of healthy adults receiving heterologous or homologous booster COVID-19 vaccine after a single prime dose of Ad26.COV2.S in a phase II randomised open-label trial, BaSiS

**DOI:** 10.1101/2024.02.19.24303026

**Authors:** Faeezah Patel, Jean Le Roux, Shobna Sawry, Robert Kieser, Mrinmayee Dhar, Katherine Gill, Erica Lazarus, Anusha Nana, Nigel Garrett, Penny L Moore, Alex Sigal, Glenda Gray, Helen V Rees, Barry Frank Jacobson, Lee Fairlie

## Abstract

BaSiS (**B**ooster **A**fter **Si**sonke **S**tudy) is a prospectively enrolled open-label trial in which healthy adults, with controlled co-morbidities and no prior thrombosis, who received a single Ad26.COV2.S prime vaccination primarily through the Sisonke phase IIIB open label implementation study in South Africa. An exploratory objective evaluated the clotting profiles of participants who were enrolled across 4 sites in South Africa and randomised 1:1:1:1 to receive one of full-dose Ad26.COV2.S, half-dose Ad26.COV2.S, full-dose Comirnaty or half-dose Comirnaty booster. D-dimer testing (INNOVANCE®D-Dimer Assay), as a coagulopathy marker, was conducted pre-booster (baseline) and 2 weeks post-booster. The median age among 285 participants was 42.2 years (IQR:35.5-48.7), 235/285 (82.5%) were female, 269/285 (94.4%) were Black African. Of the 40.4% (115/285) people living with HIV (PLHIV), 79.1% (91/115) were well-controlled on antiretroviral therapy. At baseline, 39.3% (112/285) had elevated d-dimers; all asymptomatic. Females and obese participants were significantly more likely to have elevated baseline d-dimers (OR=4.17; 95% CI:1.88 to 9.26 and OR=2.64; 95% CI:1.57 to 4.43, respectively). Of 169 with normal baseline d-dimers, 29 (17.2%) became elevated 2 weeks post-booster: median increase 0.23µg/ml (IQR:0.15-0.42); those receiving full-dose Comirnaty exhibited lower risk of d-dimer elevation post vaccination, compared to other booster vaccination arms (OR:0.26; 95% CI:0.07 to 0.98). PLHIV experienced significantly higher median increases compared to HIV uninfected participants (0.43 vs 0.17, p=0.004). Elevated d-dimers in asymptomatic, low-risk adults were unexpectedly common but were not associated with thromboembolism, supporting the rationale of using d-dimers only if clinically indicated. Trial Registration: South African Clinical Trails Register number DOH-27-012022-7841.

## Introduction

SARS-CoV-2, first identified in December 2019, soon resulted in a worldwide COVID-19 pandemic. Symptoms vary from asymptomatic to severe life-threatening conditions, affecting multiple organ systems and has resulted in large numbers of deaths. By April 2023, just prior to the World health Organization (WHO) announcement of the end of COVID-19 as a public health emergency, WHO estimated that over 765 million people worldwide had been infected with COVID-19 and over 6,9 million people had died (1).

The rapid development, testing and rollout of multiple vaccines during this pandemic contributed to limiting the spread of the virus and disease severity, especially in high-risk individuals (2–6). COVID-19 vaccines have, however, not been without controversy or complications. With the withdrawal of Vaxzevria (AstraZeneca, Oxford, UK, ChAdOx vaccine) in January 2021 from the South African national COVID-19 vaccine rollout after early results showed limited effectiveness against the then-circulating COVID-19 beta (B.1.315) variant of concern, alternatives were sought. In the absence of a national vaccination programme, the Sisonke phase 3B open label implementation study (ClinicalTrials.gov number NCT048387950) was, at the time, the only access to a COVID-19 vaccine in South Africa.

A single Janssen Ad26.COV2.S (hereafter referred to as Ad26.COV2.S) COVID-19 vaccine was provided via Sisonke, exclusively to Health Care Workers (HCW) between 17 Feb 2021 and 17 May 2021. Subsequently, the South African National Department of Health (SA NDoH) rolled out the national vaccination programme with Ad26.COV2.S and vaccine access was expanded in a stepwise manner to include either the two-dose Pfizer Comirnaty (Pfizer BNT162b2/Pfizer-BioNTech) vaccine (hereafter referred to as Comirnaty) or a single dose Ad26.COV2.S in adults and adolescents. Booster vaccinations were offered late December 2021 and since February 2022, homologous and heterologous booster COVID-19 vaccines have been made available.

In April 2021, Ad26.COV2.S was linked to a thromboembolic syndrome later named Thrombosis with Thrombocytopaenia Syndrome (TTS) (7), which is usually seen in the first two weeks post vaccination. TTS is defined as thrombosis in an unusual location (ie, cerebral vein, visceral artery or vein, extremity artery, central artery or vein) and new-onset thrombocytopenia (ie, platelet count <150 x 10^9^/L) OR new-onset thrombocytopaenia (ie, platelet count < 150 x10^9^/L), thrombosis in an extremity vein or pulmonary artery in the absence of thrombosis at an unusual location, and a positive anti-PF4 antibody enzyme linked immunosorbent assays (ELISA) test or functional Heparin-Induced Thrombocytopenia (HIT) platelet test occurring any time after vaccination (8).

Globally, approximately 4 cases of TTS per million vaccinations have been reported in males and females of all ages but most commonly in females aged 30-49 years (1 case per 100,000 vaccinations). Approximately 15% of TTS cases are fatal (9). Similar incidence was seen in the South African Sisonke study: in the 477 234 participants there were 29 thrombotic events of which only 2 were determined to be TTS (0.45 events per 100 000 population) (10).

Despite the FDA recommending a preference for mRNA vaccines since May 2022 due to TTS concerns, Ad26.COV2.S is still used, mainly in resource-limited settings, and has been proven in clinical trials and real-world situations to prevent severe SARS-CoV-2 infection (11). Also, due to the infrequency of TTS, the risk benefit ratio still favours the use of Ad26.COV2.S where vaccine options are limited (12).

D-dimer is a fibrin degradation product produced after fibrinolysis of blood clots and, as such, d-dimer testing can be useful in the diagnosis of TTS and other thrombotic events. While d-dimer tests are generally sensitive, their specificity for thromboembolism is poor and d-dimers may be elevated purely due to inflammation, in pregnancy, trauma, burns, infections, cancer, massive bleeding, HIV at seroconversion and in chronic disease, cardiovascular disease, liver and renal disease, surgery, and physiological increases with age (13,14). Presence of these conditions often result in higher levels of d-dimer in an otherwise clinically-well individual (15). Thromboembolism itself is not uncommon and the risks increase with age, comorbidities such as cardiovascular disease and metabolic syndromes, lifestyle factors especially smoking and alcohol use, and living with HIV (16).

In COVID-19 infected individuals, d-dimers may serve as a marker of COVID-19 severity, prognosis and mortality, resulting in the increased use of this test worldwide (17). Here we report on the early clotting profiles among participants from the BaSiS study, which evaluated safety and immunogenicity responses in people previously vaccinated with a prime Ad26.COV2.S vaccine, then randomised to a full or half-dose Ad26.COV2.S or Comirnaty booster vaccine, in whom d-dimers were measured at baseline (pre-booster) and at 2 weeks post-booster vaccination.

## Materials and Methods

### Study design

**B**ooster **A**fter **Si**sonke **S**tudy (BaSiS) is an ongoing phase II randomised open-label trial to compare the cellular and humoral responses of four SARS-CoV-2 booster vaccine regimens in participants who originally received a single dose of the Ad26.COV2.S vaccine through the Sisonke phase IIIB implementation study or via the SA NDoH rollout (18). The four booster vaccination regimens are full-dose Ad26.COV2.S (5x 10^10^ vp/ml, 0.25 ml), half-dose Ad26.COV2.S (2.6x 10^10^ vp/ml, 0.13 ml), full-dose Comirnaty vaccine (30mcg) and half-dose Comirnaty vaccine (15mcg). BaSiS, currently in the follow-up phase, is being conducted at four research sites in South Africa, Wits RHI Shandukani Research Centre and the Perinatal HIV Research Unit (PHRU) in Johannesburg, CAPRISA eThekwini Clinical Research Site in Durban and Desmond Tutu Health Foundation Masiphumelele Clinical Research Site in Cape Town.

The BaSiS study was approved by Wits University’s Human Research Ethics Committee (Reference no 211001B), Biomedical Research Ethics Committee (Reference no BREC/00003487/2021), University of Cape Town (UCT) Faculty of Health Sciences Human Research Ethics Committee (Reference no 680/2021) and the South African Health Products Regulatory Authority (SAHPRA Reference no 20210423). It was registered on the South African Clinical Trails Register (DOH-27-012022-7841), with received approval on 25 Jan 2022, reflecting the actual trial start date of 08 Dec 2021, in keeping with the start of enrolment initiation to the study. Trial safety monitoring was overseen by the Sisonke PSRT, and monitoring was performed by the Hutchinson Centre Research Institute of South Africa (HCRISA).

The primary objectives of BaSiS include evaluation of immunogenicity, safety and reactogenicity of the four booster vaccination regimens, stratified by age and HIV status. These primary outcomes are not included in this analysis and will be published separately. Here, we report the results from an exploratory objective evaluating the clotting profiles of participants by booster vaccination arm, age, and HIV status.

### Procedures

Enrolment took place between 08 Dec 2021 – 28 Jul 2022. Participants were vaccinated at the enrolment (baseline) visit and then had follow-up visits at 2 weeks, 3 months, and 6 months post-booster vaccination.

Full blood count (FBC) and d-dimer levels were tested at baseline (pre-vaccination) and at week 2. Interim visits were conducted for safety-related concerns including abnormal blood results. D-dimer tests, classified into ELISA, latex-enhanced agglutination assays and immunofluorescent assays (14,19), have different sensitivities, specificities and cut-off values. Results can be reported in a variety of measurement units and in D-dimer units (DDU) or fibrinogen equivalent units (FEU). Sample collection and processing, lack of standardization across assay types, differences in thresholds for age and comorbid conditions can affect d-dimer results and there is no International Reference Standard (14,20). We tested all samples at Bio Analytical Research Corporation South Africa (BARC SA) using the INNOVANCE® D-Dimer assay, a particle-enhanced immunoturbidimetric (immune-based) assay (21,22) which has previously been compared to a highly validated ELISA assay, VIDAS D-dimer Exclusion (bioMerieux Inc. Durham, North Carolina, USA), and demonstrated strong correlation and good consistency between assays [Deep Vein Thrombosis sensitivity 99% (95% confidence intervals (CI): 97% to 99%) vs 100% (95% CI: 82% to 100%) and Deep Vein Thrombosis specificity 40% [(95% CI: 38% to 40%) vs 42% (95% CI: 37% to 46%) respectively] (14). Since d-dimer levels increase physiologically with age, an American College of Emergency Physicians age-related adjustment was used in individuals >50 years of age. Also, an INNOVANCE® D-Dimer assay validation study showed that this adjustment increased specificity with only a 1% loss of sensitivity (23). For these participants, the formula: age in years x 10 ug/L (converted as age in years x 0.01 µg/ml based on the units used in our study), was used for interpreting d-dimer results and determining elevations (ie a d-dimer value higher than the age-adjusted value was considered elevated) (24).

Participants with raised d-dimers were monitored more frequently and discussed with the Haematology Department at Charlotte Maxeke Johannesburg Academic Hospital and on weekly Protocol Safety Review Team (PSRT) meetings. The PSRT included medical specialists in the fields of internal medicine, infectious diseases, haematology and pulmonology, as well as members of the Sisonke protocol and safety teams.

### Participants

Initially, health care workers over 30 years of age, who received a single prime Ad26.COV2.S during the Sisonke study, without co-morbidities or with well controlled co-morbidities, including diabetes mellitus, hypertension and clinically well people living with HIV (PLHIV) regardless of viral load and CD4 count, were enrolled. A protocol amendment implemented on 28 Apr 2022 amended the eligibility criteria to reduce the age of eligibility to 18 and to include those who received the single prime dose of Ad26.COV2.S through the SA NDoH rollout. Women of childbearing potential were not expected to use contraception; however, a negative pregnancy test was required at vaccination. Those with previous SARS-CoV-2 infection > 28 days prior to enrolment were included if clinically well. A history of previous severe allergic reaction and any thromboembolic disease were exclusionary. Body mass index (BMI) measurements were recorded but were not an eligibility criterion.

### Sample size

The main study was powered to detect at least a 6 to 7.7-fold increase in antibody-mediated neutralisation after receiving the half-dose Comirnaty vaccine, with 90% probability. The study aimed to enrol 300 participants, with at least 10% being over the age of 55 and at least a third being PLHIV. This secondary analysis included all participants from the main study.

### Randomisation

Participants were individually randomised in a 1:1:1:1 ratio to one of four arms: full-dose Ad26.COV2.S (5x 10^10^ vp/ml, 0.25 ml); half-dose Ad26.COV2.S (2.6x 10^10^ vp/ ml, 0.13 ml); full-dose Comirnaty vaccine (30mcg); half-dose Comirnaty vaccine (15mcg). Randomisation was performed in masked block sizes of 4 to16, stratified by study site and HIV status, with a 2:1 ratio of HIV uninfected people to PLHIV in each arm. Randomisation lists were uploaded to the study REDCap (Research Electronic Data Capture) database by the data manager for electronic implementation; once the study site investigator entered the HIV status of the participant into the REDCap Demographic Form, REDCap applied the random allocation table and assigned the participant to a study arm.

### Statistical analysis

Medians and interquartile ranges (IQR) were calculated for continuous data with a non-normal distribution. Categorical variables were described using proportions and 95% confidence interval (CI). Elevated d-dimer was defined as d-dimer > 0.5 µg/ml for persons up to 50 years of age (22), age-related cut-offs were used for persons over the age of 50 years, as previously described. For the analyses of the entire cohort, the proportion of participants with elevated d-dimers was determined by the number of participants with elevated d-dimers divided by the total number with d-dimer results at baseline. For the analyses of those with normal d-dimers at baseline, proportions were determined by dividing the number with elevated d-dimers at week 2 by the number with normal d-dimers at baseline. Univariate logistic regression was used to explore the association between demographic and clinical factors, and elevated d-dimers at baseline.

Participants with elevated d-dimers at baseline were excluded from analyses investigating factors associated with elevated d-dimers post vaccination. Univariate and multivariable logistic regression were performed to explore factors associated with elevated d-dimers levels among participants with a normal d-dimer at baseline. In a separate analysis we explored factors associated with sustained elevated d-dimer levels among participants with an elevated d-dimer at baseline. For multivariable models, variables with significant associations to the outcome in univariate stepwise analyses, as well as factors considered clinically important according to available literature (sex, age, hypertension, obesity, and living with HIV) and by the research team, were retained in the base model. For the remaining variables, a reverse stepwise selection was carried out using a significance cut-off of 25%. Unadjusted and adjusted odds ratios, as well as associated 95% CI, were computed.

A case series, depicting relevant clinical characteristics, is presented for participants that had a normal d-dimer result at baseline and a raised d-dimer result thereafter (Supplementary Table 1).

Median d-dimer increases experienced by participants with a normal d-dimer result at baseline and an elevated d-dimer result at 2 weeks visit, were compared between groups using either the Kruskal-Wallis equality-of-populations rank test or the two-sample Wilcoxon rank-sum test, as appropriate.

The Research Electronic Data Capture (REDCap) application (25) was used for randomisation of participants to their study arms, data entry and as the data management system for the study. Analysis was carried out using Stata 15.1 (Stata Corp, College Station, TX, USA).

### Data Availability Statement

All relevant data are within the manuscript and its Supporting Information files.

## Results

Of the 289 participants enrolled onto the study and randomly assigned to one of the 4 booster vaccination arms, 285 were included in this analysis (one participant had a prior undisclosed history of thrombosis and three did not have baseline d-dimer measures). The median age was 42.2 years (IQR: 35.5 – 48.7), 235 (82.5%) were female and 269 (94.4%) were of Black African ethnicity. Just over 40% were PLHIV, of whom 20.9% either had a low CD4 count (<350 cells/µl) or an elevated HIV viral load (>40 copies/ml) (Table 1). Of the female participants, 14 (6.0%) were on oestrogen-containing contraception, 50 (21.3%) were on a non-oestrogen-containing contraceptive and 171 (72.8%) were not taking a hormonal contraceptive. Almost two-thirds (173/285) of participants, were obese (BMI ≥ 30) and 80 (28.1%) had a history of hypertension or had an elevated blood pressure at baseline (Table 1).

**Table 1:**
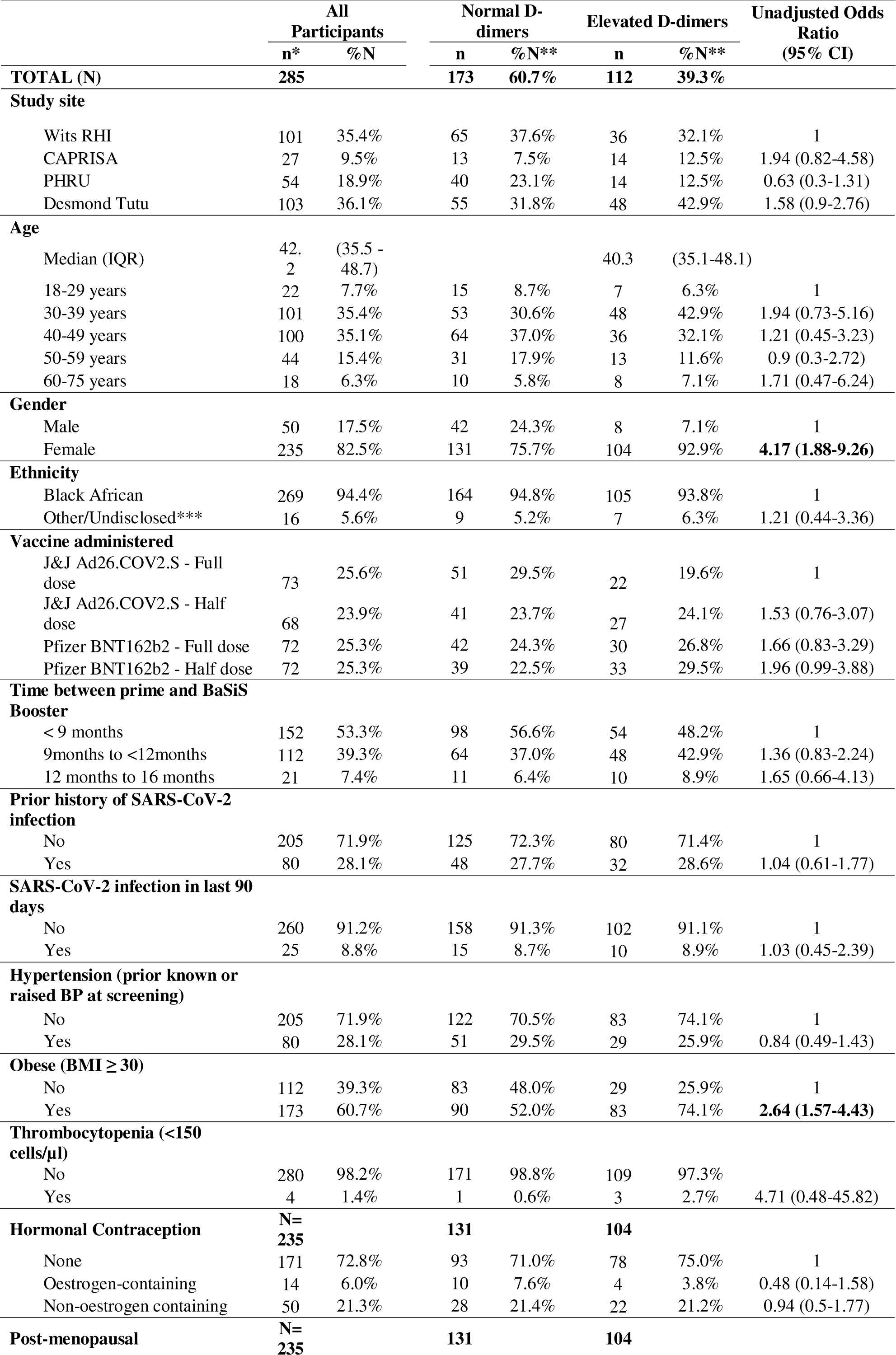
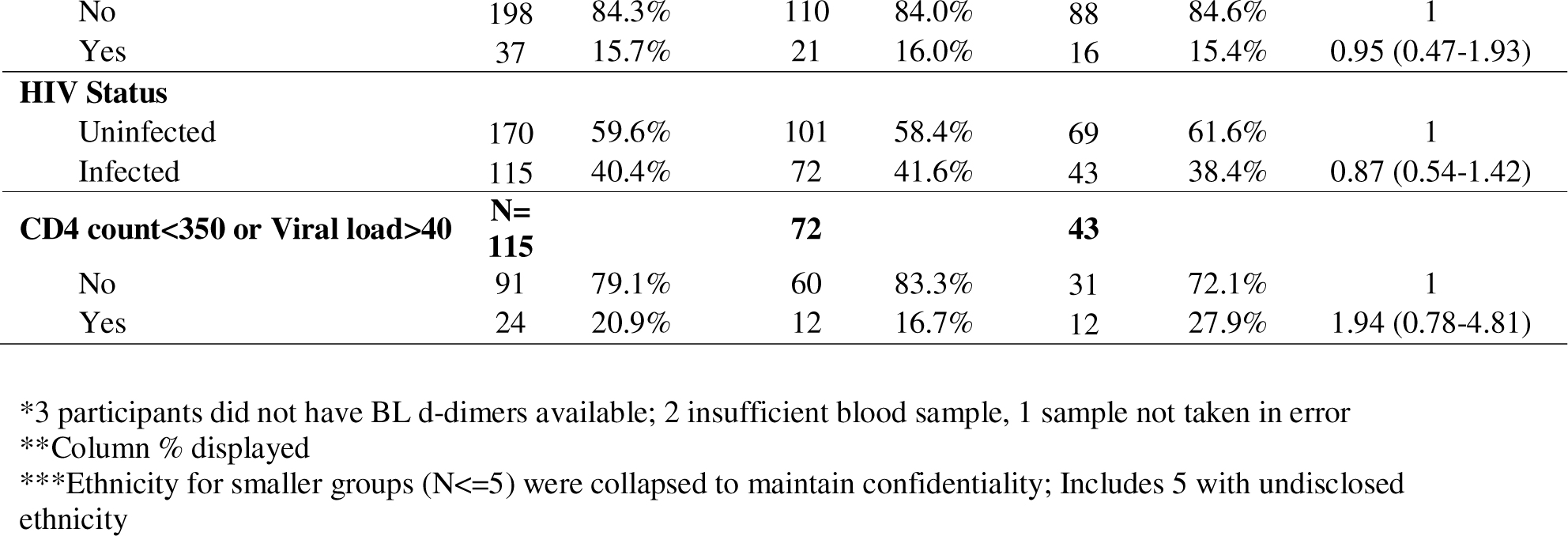
Demographic and clinical characteristics associated with elevated d-dimers at the baseline visit, prior to the administration of the booster vaccine.

At baseline, 39.3% (112/285) had elevated d-dimers, ranging between 0.50 to 12.82 µg/ml, with females and obese participants significantly more likely to have elevated d-dimers (OR=4.17; 95% CI:1.88 to 9.26 and OR=2.64; 95% CI:1.57 to 4.43, respectively). We found no associations with elevated d-dimers at baseline by age, ethnicity, hypertension, hormonal contraceptive use, menopausal status, previous reported COVID-19 infection and HIV status and poor HIV control defined as CD4+ < 350 cells/µl and HIV VL >40 copies/ml (Table 1).

Of the 112 participants with elevated d-dimers at baseline, 80 (71.4%) remained elevated, while 27 (24.1%) dropped to normal levels, and five (4.5%) did not have d-dimer results at the week 2 visit (Fig 1). Among the 173 participants with normal d-dimers at baseline, 140 (80.9%) remained normal and 29 (16.8%) became elevated, while four (2.3%) had no d-dimer result available at the week 2 visit. At the week 2 visit, 276 participants had d-dimer results available, of which 109 (39.5%) were elevated. The median change in d-dimers from baseline to week 2 in the total population of 285 was 0.00 µg/ml (IQR: −0.09-0.14). The median change in d-dimers in the 29 participants from normal baseline to an elevation at week 2 was 0.23 µg/ml (IQR: 0.05-8.28). None of the 285 participants presented with clinical evidence of thrombosis throughout the study period.

**Figure 1:**
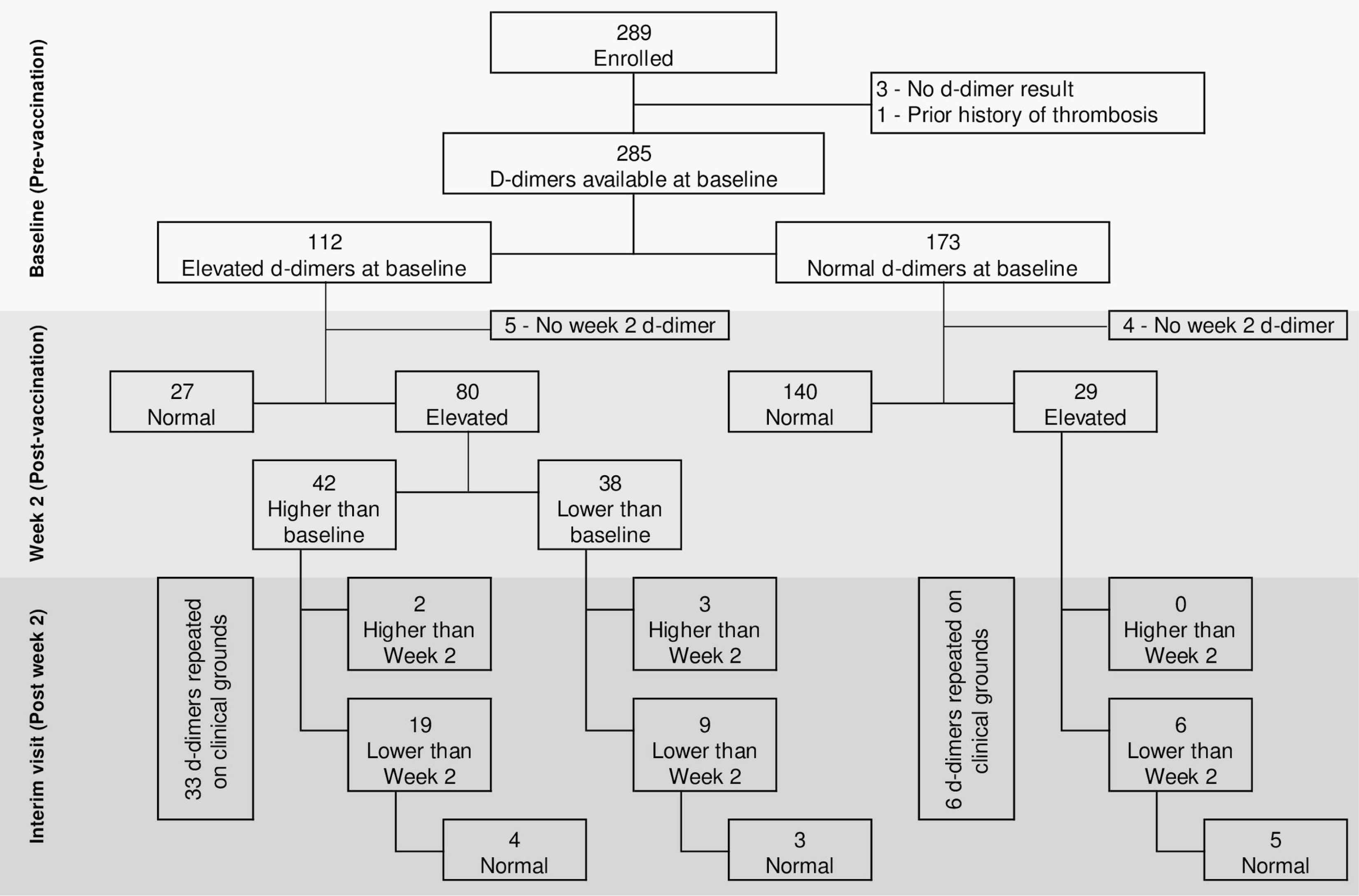
BaSiS study cohort d-dimer profile among 289 participants reflecting d-dimers at baseline, 2 weeks post-booster vaccination, and up to 4-weeks post-booster vaccination.

### Participants with normal d-dimers at baseline: factors associated with newly raised d-dimers at week 2

Among the 173 participants with normal d-dimers at baseline, 169 had a week 2 d-dimer result available; 29/169 (17.2%) were elevated. On univariate analysis of these 169 participants, females had higher odds of an elevated d-dimer at week 2, (OR:5.03; 95% CI:1.14 to 22.18) and participants who received the full-dose Comirnaty had a lower odds of d-dimer elevation at week 2, compared to full-dose Ad26.COV2.S (OR: 0.26; 95% CI: 0.07 to 0.98). We found no significant differences by site, age, HIV status or control, hypertension, obesity, hormonal contraceptive use, prior SARS-CoV-2 infection, and time between prime and booster vaccination (Table 2). On multivariable analysis, female participants had a significantly higher odds of having elevated d-dimers at week 2 (OR: 4.97; 95% CI: 1.07 to 23.18). The significant protective association found with the full-dose Comirnaty as compared to the full-dose Ad26.COV2.S did not persist in the multivariable analysis (OR: 0.26; 95% CI 0.06 to 1.06). Participant age, vaccine dose and type, hormonal contraception use and comorbid conditions were not associated with new onset d-dimer elevations at week 2 (Table 2).

**Table 2:** Demographic and clinical characteristics associated with elevated d-dimers 2 weeks post-booster vaccination, among 169 participants who had normal d-dimers at baseline.

|  | Normal d-dimers at baseline | Elevated d-dimers 2 weeks post-booster vaccination |  | Univariate analyses | Multivariable analyses |
| --- | --- | --- | --- | --- | --- |
|  | N* | n | %N | Unadjusted OR (95% CI) | Adjusted OR (95% CI) |
| **TOTAL (N)** | **169** | **29** | **17.2%** |  |  |
| **Study site** |  |  |  |  |  |
| Wits RHI | 64 | 8 | 12.5% | 1 |  |
| CAPRISA | 12 | 2 | 16.7% | 1.4 (0.26-7.58) |  |
| PHRU | 38 | 7 | 18.4% | 1.58 (0.52-4.77) |  |
| Desmond Tutu | 55 | 12 | 21.8% | 1.95 (0.73-5.2) |  |
| **Age groups** |  |  |  |  |  |
| 18-29 years | 14 | 2 | 14.3% | 1 | 1 |
| 30-39 years | 52 | 13 | 25.0% | 2 (0.39-10.14) | 2.18 (0.37-12.94) |
| 40-49 years | 62 | 9 | 14.5% | 1.02 (0.19-5.33) | 1.01 (0.16-6.51) |
| 50-59 years | 31 | 4 | 12.9% | 0.89 (0.14-5.53) | 0.83 (0.11-6.31) |
| 60-75 years | 10 | 1 | 10.0% | 0.67 (0.05-8.55) | 0.64 (0.04-10.61) |
| **Gender** |  |  |  |  |  |
| Male | 40 | 2 | 5.0% | 1 | 1 |
| Female | 129 | 27 | 20.9% | **5.03 (1.14-22.18)** | **4.97 (1.07-23.18)** |
| **Ethnicity** |  |  |  |  |  |
| Black African | 160 | 29 | 18.1% | 1 |  |
| Other/Undisclosed** | 9 | 0 | 0.0% | 1 (0-0) |  |
| **Booster Vaccine type & dose administered** |  |  |  |  |  |
| J&J Ad26.COV2.S – Full-dose | 51 | 12 | 23.5% | 1 | 1 |
| J&J Ad26.COV2.S – Half-dose | 40 | 5 | 12.5% | 0.46 (0.15-1.45) | 0.33 (0.10 – 1.12) |
| Pfizer BNT162b2 – Full-dose | 41 | 3 | 7.3% | **0.26 (0.07 – 0.98)** | 0.26 (0.06 – 1.06) |
| Pfizer BNT162b2 – Half-dose | 37 | 9 | 24.3% | 1.04 (0.39-2.81) | 1.30 (0.44 – 3.83) |
| **Booster Vaccine type administered** |  |  |  |  |  |
| J&J Ad26.COV2.S | 91 | 17 | 18.7% | 1 |  |
| Pfizer BNT162b2 | 78 | 12 | 15.4% | 0.79 (0.35-1.78) |  |
| **Time between prime and BaSiS** |  |  |  |  |  |
| **Booster vaccination** |  |  |  |  |  |
| < 9 months | 94 | 18 | 19.1% | 1 |  |
| 9 months to < 12 months | 64 | 9 | 14.1% | 0.69 (0.29-1.65) |  |
| 12 months to 16 months | 11 | 2 | 18.2% | 0.94 (0.19-4.72) |  |
| **Prior history of SARS-CoV-2 infection** |  |  |  |  |  |
| No | 123 | 24 | 19.5% | 1 |  |
| Yes | 46 | 5 | 10.9% | 0.5 (0.18-1.41) |  |
| **SARS-CoV-2 infection in last 90 days** |  |  |  |  |  |
| No | 154 | 28 | 18.2% | 1 |  |
| Yes | 15 | 1 | 6.7% | 0.32 (0.04-2.55) |  |
| **Hypertension (prior known or raised BP at screening)** |  |  |  |  |  |
| No | 118 | 23 | 19.5% | 1 | 1 |
| Yes | 51 | 6 | 11.8% | 0.55 (0.21-1.45) | 0.45 (0.15 – 1.36) |
| **Obese (BMI ≥ 30)** |  |  |  |  |  |
| No | 80 | 10 | 12.5% | 1 | 1 |
| Yes | 89 | 19 | 21.3% | 1.9 (0.82-4.38) | 2.02 (0.78 – 5.23) |
| **Thrombocytopenia (<150 x 10<sup>9</sup> cells/L)** |  |  |  |  |  |
| No | 167 | 29 | 17.4% | 1 |  |
| Yes | 1 | 0 | 0.0% | 1 (0-0) |  |
| **Hormonal Contraception** |  |  |  |  |  |
|  | N=129 |  |  |  |  |
| None | 92 | 17 | 18.5% | 1 |  |
| Oestrogen-containing | 10 | 2 | 20.0% | 1.1 (0.21-5.67) |  |
| Non-oestrogen containing | 27 | 8 | 29.6% | 1.86 (0.7-4.95) |  |
| **Post-menopausal** | N=129 |  |  |  |  |
| No | 108 | 25 | 23.1% | 1 |  |
| Yes | 21 | 2 | 9.5% | 0.35 (0.08-1.6) |  |
| **HIV Status** |  |  |  |  |  |
| Uninfected | 99 | 16 | 16.2% | 1 | 1 |
| Infected | 70 | 13 | 18.6% | 1.18 (0.53-2.65) | 1.06 (0.43 – 2.61) |
| **CD4 count<350 or Viral load>40** | N=70 |  |  |  |  |
| No | 59 | 10 | 16.9% | 1 |  |
| Yes | 11 | 3 | 27.3% | 1.84 (0.41-8.16) |  |

\*4/173 with non-elevated d-dimers at enrolment, did not have 2week d-dimer data and are excluded from this table; 1 early withdrawal, 3 missed week 2 visits
\*\*Ethnicity for smaller groups (N<=5) were collapsed to maintain confidentiality; Includes 1 with undisclosed ethnicity

### Participants with normal baseline and raised week 2 d-dimers: factors associated with higher median increase in d-dimer levels

Of the 29 participants with a new onset of elevated d-dimers at the week 2 visit, 13 were PLHIV, three of whom had poorly controlled HIV (Table 2). Four participants had d-dimer results >1.0 µg/ml and two others experienced a doubling of d-dimer values when compared to baseline. These six participants had interim visits to repeat d-dimer tests, where two remained elevated and four dropped to below 0.5 µg/ml. While HIV status was not significantly associated with an elevation of d-dimers at week 2 (Table 2), among all those who experienced an elevation, the median absolute increase in d-dimer levels was significantly higher among PLHIV (median increase: 0.43 µg/ml; IQR: 0.29-0.75) compared to HIV uninfected participants (median increase: 0.17 µg/ml; IQR: 0.11-0.24) (p=0.004) (Table 3). There were no significant differences in median increases in d-dimers for other co-variates (Table 3).

**Table 3:**
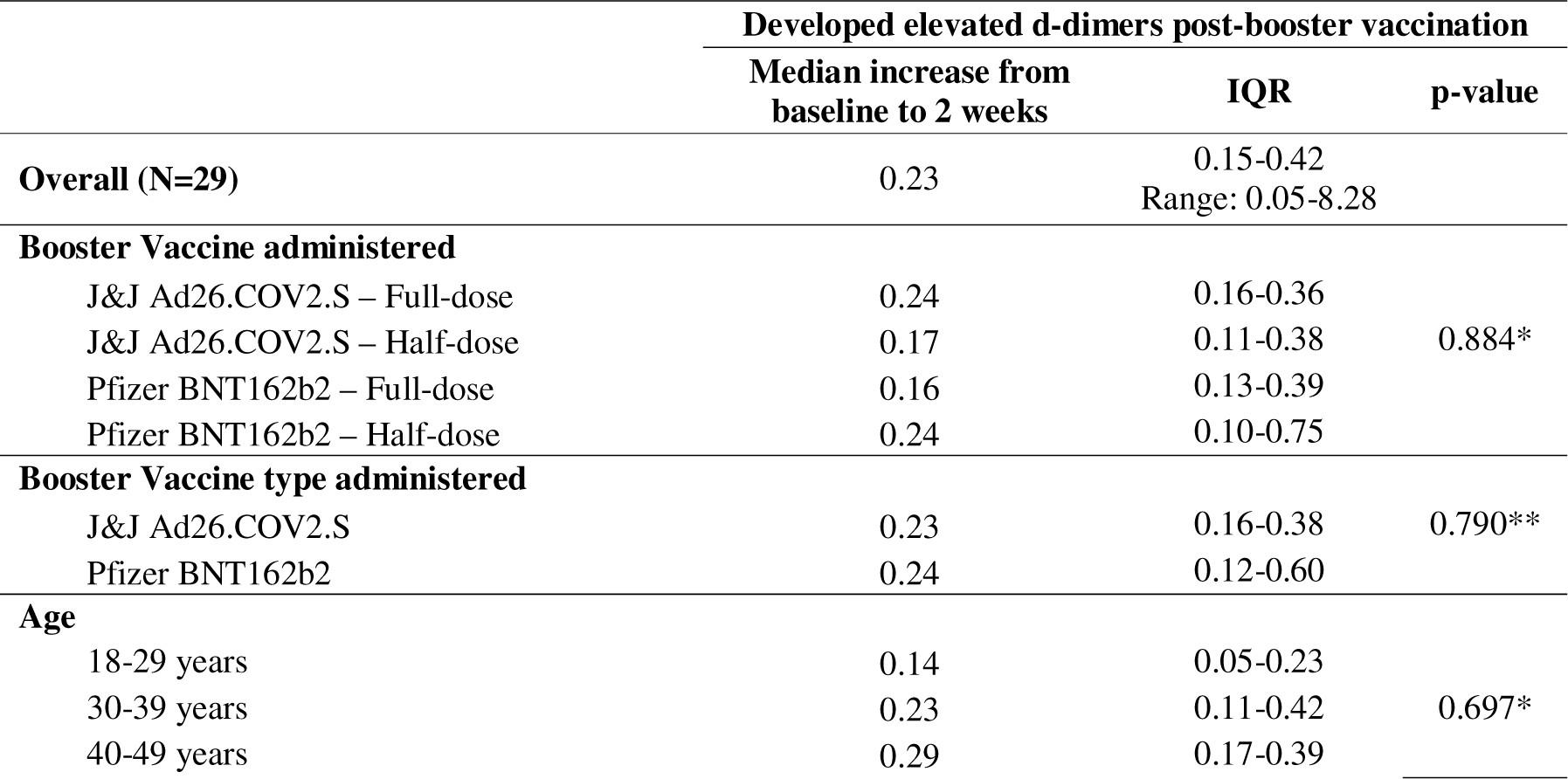
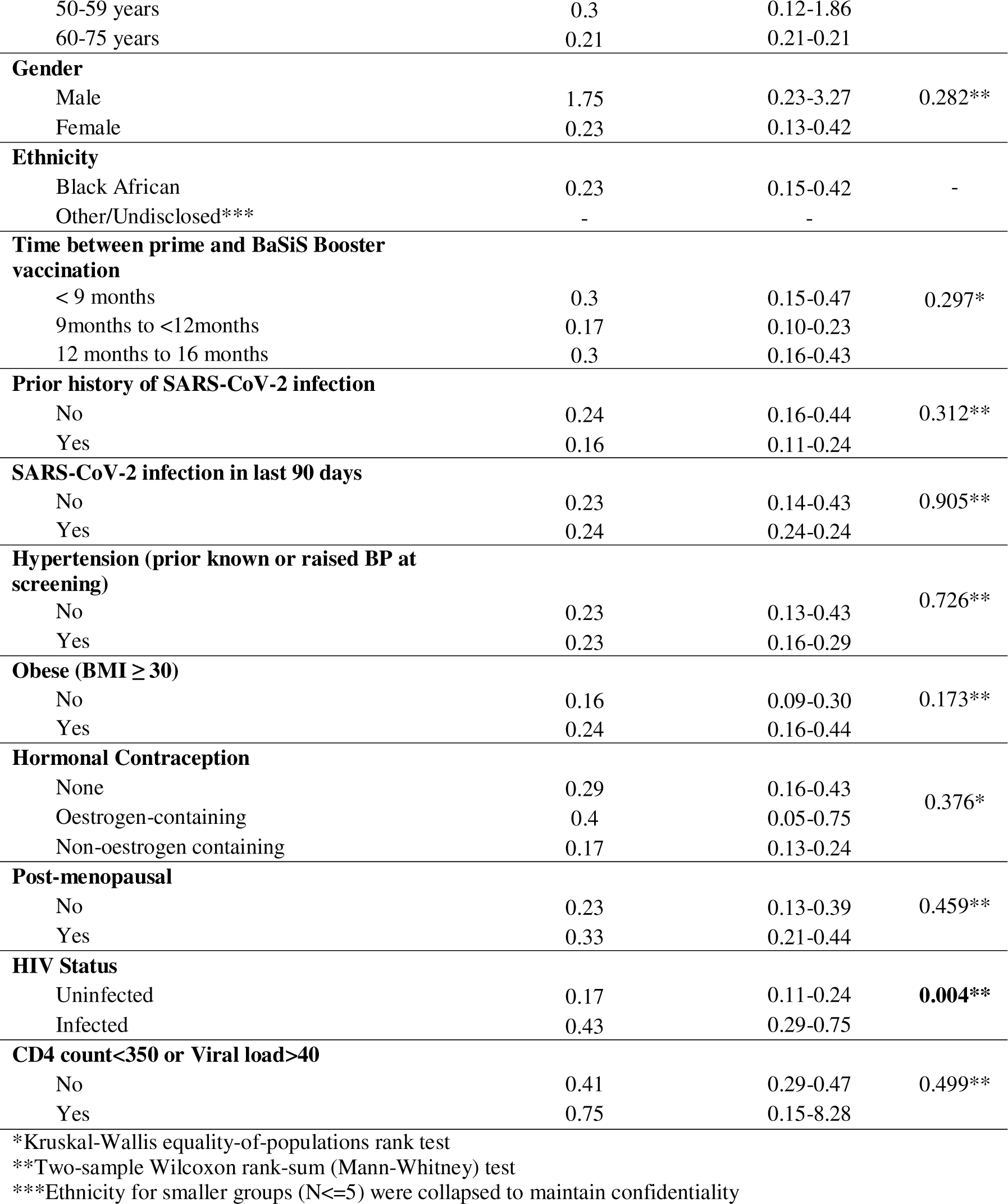
Median increases in d-dimers among the 29 participants who developed elevated d-dimers 2 weeks post-booster vaccination, stratified by demographic and clinical co-variates.

### Participants with elevated d-dimers at baseline: factors associated with sustained elevation at week 2

Among the 107 participants who had elevated d-dimers at baseline and a week 2 result available, 80 (74.8%) had sustained elevations at week 2. On both univariate and multivariable analysis, we found no significant differences by any co-variates (Supplementary Table 2). Four participants had a baseline platelet count less than 150 x 10^9^/L and three had associated increased baseline d-dimers. Two of the three maintained the thrombocytopaenia and increased d-dimers at week 2, neither were symptomatic for TTS.

## Discussion

In this exploratory analysis of the clotting profiles of participants in the BaSiS study, we found a high proportion (39.3%) of participants with elevated d-dimers prior to receipt of their allocated booster vaccination. In addition, 17.2% of participants with normal d-dimers at baseline, developed elevated d-dimers 2 weeks post-booster vaccination. None had symptoms of thrombosis pre- or post-booster vaccination and no TTS was diagnosed. Female participants and those who were obese (BMI > 30 had a significantly higher odds of having elevated d-dimers 2 weeks post-booster vaccination (OR: 4.97; 95% CI:1.07 to 23.18 and OR: 2.64; 95% CI:1.57 to 4.43 respectively), though with wide confidence intervals, possibly reflective of the low number of male participants with new onset d-dimer elevations at 2 weeks post-booster vaccination. The interpretation of these elevated baseline d-dimers is challenging as there is little data available globally on baseline levels within a clinically well population, and especially not in resource-limited settings like South Africa. We found no further associations between age, ethnicity, booster vaccine type and dose, prior COVID infection, hormonal contraception use, obesity, hypertension and HIV infection with new onset d-dimer elevations 2 weeks post-booster vaccination.

D-dimer tests support diagnoses of deep vein thrombosis and pulmonary embolism when clinical signs and symptoms are present and supported by radiological and sonar imaging, but has limited use when thromboembolism is not suspected. Its utility as a stand-alone diagnostic test is diminished because there are a number of conditions that increase d-dimer levels without the presence of thromboembolism including pregnancy, inflammation, cancer, trauma, surgery, obesity and associated cardiovascular and insulin-resistant conditions, HIV and the physiological increases that occur with age (26–28). Gender differences have been noted when screening for thromboembolism with females being more likely to have a d-dimer value above the upper limit cut-off. Currently there is no gender specific d-dimer reference range (29). Among PLHIV, data from both resource-rich and poor settings has demonstrated raised d-dimers during acute seroconversion and in chronic HIV infection (28,30). ART initiation with subsequent virological and immunological control results in a reduction of d-dimers (31–34), however, despite viral suppression, higher d-dimer levels may persist in PLHIV (35,36). To our knowledge there is no available published evidence of the effect of HIV on d-dimer results post COVID-19 vaccinations. Since South Africa still remains one of the highest HIV-burden countries globally, with almost 8 million people living with HIV, having elevated d-dimers in this population could place these individuals at higher risk of a thromboembolism.

The Ad26.COV2.S vaccine clotting concerns and that raised d-dimer results are useful in predicting COVID-19 illness severity and associated thromboembolism, and has a high predictive value for serious disease in those clinically unstable (37,38), led us to measure d-dimer evaluations as screening tests for coagulopathy risk in our study. A similar prospective observational study conducted in Croatia (n=171), compared d-dimers between Comirnaty (n=87) and Ad26.COV2.S (n=84) pre-dose and at multiple time points post-dose. This study had a similarly high proportion of female participants with 59% and 48% in the Comirnaty and Ad26.COV2.S groups, respectively. Using the INNOVANCE® D-Dimer Assay, they reported a statistically significant (P=0.004) rise in d-dimers post vaccination in both vaccine groups without clinical evidence of thromboembolism. No further sub-group analyses were performed to elucidate differences in sex and HIV status of participants (39). TREASURE reported no differences in median d-dimer values 8-10 days post vaccination (p=0.950) between the different vaccines, though with smaller representation of Ad26COV2.S and Comirnaty in the sample (40). A Norwegian study compared health care personnel who received Vaxzevria (n=521) to a control group who received no SARS-CoV-2 vaccination and had no prior history of COVID-19 infection (n=102) (41). Females constituted 76% of the Vaxzevria group and 50% of controls. Interestingly, the control group had higher mean d-dimers post vaccination (227ng/mL, 206-248) than the Vaxzevria recipients (214ng/mL, 205-224).

Smaller studies conducted in Singapore (n=18) and Italy (n=30) compared pre- and post-booster vaccination d-dimers using different assays, in participants vaccinated with two doses of Comirnaty vaccine (42,43). Median baseline d-dimers in the predominantly young (median age 35 years; IQR: 31-44) female (78%) Singaporean cohort was 0.32 µg/ml (IQR 0.22-0.38), with similar results post first (0.21 µg/ml; IQR: 0.14-0.35) and second vaccine doses (0.29 µg/ml: IQR 0.24-0.34) (p=0.35). The Italian study, with no age or gender disaggregation, reported a median baseline d-dimer of 273 ng/ml (IQR 175-360), and 214 ng/ml (IQR: 144-431) and 286ng/ml (IQR: 160-514) post first and second dose, respectively (p=0.633 baseline compared to 14 days post second dose). Their preliminary data concluded that administration of Comirnaty vaccines did not result in a hypercoagulable state. Neither study provided descriptive data about the BMI or HIV status of the participants. Contrary to our results, among 30 Sudanese participants previously vaccinated with Ad26COV2.S, majority of whom were female (64.7%), who received a Ad26COV2.S booster on study, a statistically significant increase in mean d-dimers (0.17±0.03 vs 0.37±0.08, p-value ≤ 0.05) was noted post-booster vaccination. No symptoms of thrombosis, and no difference in mean d-dimers post vaccination between females and males was noted (44).

Our study enrolled PLHIV (40%), many clinically well controlled and found no significant differences in elevated d-dimers at baseline compared to 2 weeks post-booster vaccination. This is likely due to many having suppressed HIV viral loads or high CD4 cell counts. However, in PLHIV who had an elevated d-dimer at 2 weeks post-booster vaccination, the absolute median increase in d-dimers was higher compared to HIV uninfected people who had d-dimer elevation. The clinical relevance of this is uncertain, as the increase in d-dimer level was very small.

This sub-analysis of the BaSiS study had a few limitations. The study was not adequately powered to evaluate d-dimer increases as evidenced by the wide confidence intervals around the point estimates, resulting in a lack of precision. Despite this, it is possibly the largest cohort of its kind, describing d-dimer elevations among those who received a Ad26COV2.S prime vaccination followed by a booster vaccination. We found an association between being female and obese with elevated baseline d-dimers. The female predominance in the cohort may present a selection bias, but is reflective of the population of health care workers who are predominantly female. (45). The high proportion of obese participants (60.7%) in our study is attributed to the study’s female predominance but is also reflective of the obesity rates in the general South African female population (67%) (46). TTS diagnosis requires anti-platelet factor 4 testing and this was not measured in our study, though none of our participants had symptoms of thrombosis while on study. Despite these limitations, the data represented here contributes to the limited research published and strengthens the advocacy for more conservative use of d-dimers in healthy asymptomatic patients in the context of COVID-19 vaccination.

## Conclusions

In the BaSiS study, a cohort consisting mostly of females with BMIs > 30 kg/m^2^, a high proportion had elevated baseline d-dimers and about 17% had an increase in d-dimers from normal baseline post vaccination. We found no association between any booster vaccine type or dose with raised d-dimers post-booster vaccination, but people who received a Cominarty full dose booster, had lower odds of d-dimer elevation post-booster. While HIV status was not associated with an elevation of d-dimers post vaccination, among those who experienced an elevation, the median increase in d-dimers was higher among PLHIV, however the clinical relevance is not known. This data adds to the small body of evidence currently available against routine d-dimer testing in the era of COVID-19 vaccination through prospective collection of pre- and post vaccination d-dimer data especially in PLHIV.

## Supporting information

SANCTR Basis approval

Basis Protocol v2.0

## Acknowledgements

We thank all the trial participants who contributed to this study. We would like to thank the vaccine sponsor: South African National Department of Health SA NdoH for Comirnaty, and Janssen Pharmaceuticals for Janssen Ad26.COV2.S.

## Author Contributions

**Conceptualization:** FP, LF, PLM, AS, SS, KG, NG, GG, HVR, EL in collaboration with the sponsor (SAMRC). **Investigation:** FP, SS, JLR, RK, MD, KG, EL, AN, NG, PLM, AS, GG, HVR, BJ, LF. **Methodology:** FP, LF, PLM, AS, SS, KG, NG, GG, HVR, EL. **Data curation:** SS, JLR, FP, KG, EL, AN, NG, LF. **Formal analysis**: SS, JLR**. Project administration:** FP, LF, PLM, AS, SS, KG, NG, EL. **Writing (Original Draft Preparation):** FP, MD, LF, SS, JLR, RK. **Writing (Review & Editing):** All authors reviewed and approved the final draft.

