## Supplementary 1 Table 1 for "Clot Twist – D-dimer analysis of healthy adults receiving heterologous or homologous booster COVID-19 vaccine after a single prime dose of Ad26.COV2.S in a phase II randomised open-label trial, BaSiS"

**Supplementary Table 1:** Case-series of 29 participants with normal d-dimers at baseline and elevated d-dimers 2 weeks post-booster vaccination

|  | **Enrolment** | | | | | | |  | | **Week 2 visit** | | |  | | **Interim visit** | | |
| --- | --- | --- | --- | --- | --- | --- | --- | --- | --- | --- | --- | --- | --- | --- | --- | --- | --- |
| **Record ID** | **Vaccine administered*** | **HIV Status** | **CD4 count<350 or VL>40** | **Prior history of SARS-CoV-2 infection** | **Hypertension** | **Thrombocytopenia** | **D-dimer** | |  | | **Days after enrolment visit** | **D-dimer** | |  | | **Days after enrolment visit** | **D-dimer** |
| 5737-1050 | Full-dose Ad26.COV2.S | HIV infected | Yes | No | No | No | 0.44 | |  | | 11 | 0.59 | |  | |  |  |
| 5738-1009 | Half-dose Comirnaty | HIV infected | Yes | No | No | No | 0.46 | |  | | 17 | 8.74 | |  | | 23 | 4.68 |
| 5738-1010 | Half-dose Comirnaty | HIV infected | Yes | No | No | No | 0.31 | |  | | 15 | 1.06 | |  | | 38 | 0.35 |
| 5735-1013 | Full-dose Ad26.COV2.S | HIV infected | No | No | No | No | 0.45 | |  | | 16 | 0.92 | |  | | 44 | 0.66 |
| 5735-1039 | Full-dose Ad26.COV2.S | HIV infected | No | No | No | No | 0.44 | |  | | 14 | 0.61 | |  | |  |  |
| 5735-1041 | Full-dose Ad26.COV2.S | HIV infected | No | No | Yes | No | 0.44 | |  | | 16 | 0.73 | |  | |  |  |
| 5738-1031 | Full-dose Ad26.COV2.S | HIV infected | No | No | No | No | 0.36 | |  | | 13 | 0.66 | |  | |  |  |
| 5738-1114 | Full-dose Ad26.COV2.S | HIV infected | No | No | No | No | 0.35 | |  | | 14 | 0.78 | |  | |  |  |
| 5735-1015 | Half-dose Ad26.COV2.S | HIV infected | No | No | No | No | 0.29 | |  | | 13 | 0.67 | |  | | 28 | 0.31 |
| 5736-1015 | Half-dose Ad26.COV2.S | HIV infected | No | No | No | No | 0.32 | |  | | 15 | 1.31 | |  | | 29 | 0.49 |
| 5736-1021 | Half-dose Comirnaty | HIV infected | No | No | Yes | No | 0.37 | |  | | 13 | 0.81 | |  | |  |  |
| 5738-1049 | Half-dose Comirnaty | HIV infected | No | No | No | No | 0.49 | |  | | 14 | 3.76 | |  | | 24 | 0.48 |
| 5738-1065 | Half-dose Comirnaty | HIV infected | No | No | Yes | No | 0.45 | |  | | 15 | 0.54 | |  | |  |  |
| 5735-1068 | Full-dose Ad26.COV2.S | HIV uninfected |  | Yes | Yes | No | 0.46 | |  | | 14 | 0.7 | |  | |  |  |
| 5735-1110 | Full-dose Ad26.COV2.S | HIV uninfected |  | No | No | No | 0.45 | |  | | 14 | 0.68 | |  | |  |  |
| 5737-1056 | Full-dose Ad26.COV2.S | HIV uninfected |  | No | No | No | 0.48 | |  | | 11 | 0.53 | |  | |  |  |
| 5738-1058 | Full-dose Ad26.COV2.S | HIV uninfected |  | No | No | No | 0.32 | |  | | 14 | 0.74 | |  | |  |  |
| 5738-1097 | Full-dose Ad26.COV2.S | HIV uninfected |  | No | No | No | 0.46 | |  | | 11 | 0.62 | |  | |  |  |
| 5737-1031 | Full-dose Ad26.COV2.S | HIV uninfected |  | No | Yes | No | 0.62 | |  | | 14 | 0.83 | |  | |  |  |
| 5735-1030 | Half-dose Ad26.COV2.S | HIV uninfected |  | Yes | No | No | 0.39 | |  | | 14 | 0.5 | |  | |  |  |
| 5737-1026 | Half-dose Ad26.COV2.S | HIV uninfected |  | No | No | No | 0.37 | |  | | 13 | 0.54 | |  | |  |  |
| 5738-1024 | Half-dose Ad26.COV2.S | HIV uninfected |  | No | No | No | 0.4 | |  | | 20 | 0.5 | |  | |  |  |
| 5735-1042 | Full-dose Comirnaty | HIV uninfected |  | Yes | No | No | 0.29 | |  | | 15 | 0.68 | |  | |  |  |
| 5738-1026 | Full-dose Comirnaty | HIV uninfected |  | Yes | Yes | No | 0.46 | |  | | 14 | 0.62 | |  | |  |  |
| 5738-1047 | Full-dose Comirnaty | HIV uninfected |  | No | No | No | 0.44 | |  | | 11 | 0.57 | |  | |  |  |
| 5737-1008 | Half-dose Comirnaty | HIV uninfected |  | No | No | No | 0.32 | |  | | 13 | 0.55 | |  | |  |  |
| 5737-1012 | Half-dose Comirnaty | HIV uninfected |  | No | No | No | 0.48 | |  | | 17 | 0.55 | |  | |  |  |
| 5737-1022 | Half-dose Comirnaty | HIV uninfected |  | Yes | No | No | 0.44 | |  | | 13 | 0.54 | |  | |  |  |
| 5738-1061 | Half-dose Comirnaty | HIV uninfected |  | No | No | No | 0.32 | |  | | 14 | 0.56 | |  | |  |  |

^*^ Group A= J&J Ad26.COV2.S - Full dose; Group B= J&J Ad26.COV2.S - Half dose; Group C= Pfizer BNT162b2 - Full dose; Group D= Pfizer BNT162b2 - Half dose

**Supplementary Table 2:** Demographic and clinical characteristics associated with sustained elevated d-dimers 2 weeks post-booster vaccination, among 107 participants who had elevated d-dimers at baseline

|  |  | **Elevated d-dimers at baseline** | **Elevated d-dimers  2 weeks post-booster vaccination** | | **Univariate analyses** | **Multivariable analyses** |
| --- | --- | --- | --- | --- | --- | --- |
|  |  | **N*** | **n** | **%N** | **Unadjusted OR (95% CI)** | **Adjusted OR (95% CI)** |
| **TOTAL (N)** | | **107** | **80** | **74.8%** |  |  |
| **Study site** | |  |  |  |  |  |
|  | Wits RHI | 35 | 27 | 77.1% | 1 |  |
|  | CAPRISA | 12 | 10 | 83.3% | 1.48 (0.27-8.20) |  |
|  | PHRU | 13 | 10 | 76.9% | 0.99 (0.22-4.48) |  |
|  | Desmond Tutu | 47 | 33 | 70.2% | 0.70 (0.26-1.91) |  |
| **Age groups** | |  |  |  |  |  |
|  | 18-29 years | 6 | 5 | 83.3% | 1 | 1 |
|  | 30-39 years | 46 | 34 | 73.9% | 0.57 (0.06-5.35) | 0.62 (0.06-5.98) |
|  | 40-49 years | 35 | 26 | 74.3% | 0.58 (0.06-5.63) | 0.67 (0.06-6.88) |
|  | 50-59 years | 13 | 10 | 76.9% | 0.67 (0.05-8.16) | 0.88 (0.07-11.73) |
|  | 60-75 years | 7 | 5 | 71.4% | 0.50 (0.03-7.45) | 0.61 (0.04-9.72) |
| **Age groups** | |  |  |  |  |  |
|  | 18-29 years | 6 | 5 | 83.3% | 1 |  |
|  | 30-39 years | 46 | 34 | 73.9% | 0.57 (0.06-5.35) |  |
|  | 40-49 years | 35 | 26 | 74.3% | 0.58 (0.06-5.63) |  |
|  | ≥50 years | 20 | 15 | 75.0% | 0.60 (0.06-6.44) |  |
| **Gender** | |  |  |  |  |  |
|  | Male | 8 | 5 | 62.5% | 1 | 1 |
|  | Female | 99 | 75 | 75.8% | 1.88 (0.42-8.43) | 1.86 (0.33-10.6) |
| **Ethnicity** | |  |  |  |  |  |
|  | Black African | 100 | 76 | 76.0% | 1 |  |
|  | Other/Undisclosed** | 7 | 4 | 57.1% | 0.42 (0.09-2.02) |  |
| **Booster Vaccine type & dose administered** | |  |  |  |  |  |
|  | J&J Ad26.COV2.S – Full-dose | 22 | 15 | 68.2% | 1 |  |
|  | J&J Ad26.COV2.S – Half-dose | 25 | 17 | 68.0% | 0.99 (0.29-3.39) |  |
|  | Pfizer BNT162b2 – Full-dose | 30 | 24 | 80.0% | 1.87 (0.53-6.63) |  |
|  | Pfizer BNT162b2 – Half-dose | 30 | 24 | 80.0% | 1.87 (0.53-6.63) |  |
| **Booster Vaccine type administered** | |  |  |  |  |  |
|  | J&J Ad26.COV2.S | 47 | 32 | 68.1% | 1 |  |
|  | Pfizer BNT162b2 | 60 | 48 | 80.0% | 1.88 (0.78-4.53) |  |
| **Time between prime and BaSiS Booster vaccination** | |  |  |  |  |  |
|  | < 9 months | 51 | 38 | 74.5% | 1 |  |
|  | 9months to <12months | 47 | 35 | 74.5% | 1.00 (0.40-2.48) |  |
|  | 12 months to 16 months | 9 | 7 | 77.8% | 1.20 (0.22-6.51) |  |
| **Prior history of SARS-CoV-2 infection** | |  |  |  |  |  |
|  | No | 77 | 57 | 74.0% | 1 |  |
|  | Yes | 30 | 23 | 76.7% | 1.15 (0.43-3.10) |  |
| **SARS-CoV-2 infection in last 90 days** | |  |  |  |  |  |
|  | No | 98 | 73 | 74.5% | 1 |  |
|  | Yes | 9 | 7 | 77.8% | 1.20 (0.23-6.15) |  |
| **Hypertension (prior known or raised BP at screening)** | |  |  |  |  |  |
|  | No | 79 | 61 | 77.2% | 1 | 1 |
|  | Yes | 28 | 19 | 67.9% | 0.62 (0.24-1.61) | 0.69 (0.24-1.99) |
| **Obese (BMI ≥ 30)** | |  |  |  |  |  |
|  | No | 27 | 20 | 74.1% | 1 | 1 |
|  | Yes | 80 | 60 | 75.0% | 1.05 (0.39-2.85) | 1.02 (0.32-3.19) |
| **Thrombocytopenia**  **(<150 x 10^9^ /L)** | |  |  |  |  |  |
|  | No | 104 | 78 | 75.0% | 1 |  |
|  | Yes | 3 | 2 | 66.7% | 0.67 (0.06-7.66) |  |
| **Hormonal Contraception** | | N=99 |  |  |  |  |
|  | None | 74 | 56 | 75.7% | 1 |  |
|  | Oestrogen-containing | 4 | 4 | 100.0% | 1 (0-0) |  |
|  | Non-oestrogen containing | 21 | 15 | 71.4% | 0.80 (0.27-2.38) |  |
| **Post-menopausal** | | N=99 |  |  |  |  |
|  | No | 84 | 64 | 76.2% | 1 |  |
|  | Yes | 15 | 11 | 73.3% | 0.86 (0.25-3.00) |  |
| **HIV Status** | |  |  |  |  |  |
|  | Uninfected | 67 | 48 | 71.6% | 1 | 1 |
|  | Infected | 40 | 32 | 80.0% | 1.58 (0.62-4.05) | 1.70 (0.63-4.55) |
| **CD4 count<350 or Viral load>40** | | N=40 |  |  |  |  |
|  | No | 29 | 22 | 75.9% | 1 |  |
|  | Yes | 11 | 10 | 90.9% | 3.18 (0.34-29.43) |  |

*5/112 with elevated d-dimers at baseline, did not have 2w d-dimer data and are excluded from this table; 1 early withdrawal, 3 missed week 2 visits, 1 too ill for blood draw

**Ethnicity for smaller groups (N<=5) were collapsed to maintain confidentiality
