## Supplementary material for "Clot Twist – D-dimer analysis of healthy adults receiving heterologous or homologous booster COVID-19 vaccine after a single prime dose of Ad26.COV2.S in a phase II randomised open-label trial, BaSiS": SANCTR Basis approval

### South African National Clinical Trials Registry

**South African Medical Research Council, Cochrane South Africa**
**PO Box 19070, Tygerberg, 7505, South Africa**
** Website: sanctr.samrc.ac.za**

| **Trial no.:** | DOH-27-012022-7841 | **Date of Approval:** | 25/01/2022 |
|---|---|---|---|
| **Trial Status:** | Approved | | |

| **TRIAL DESCRIPTION** | |
|---|---|
| **Public title** | BaSiS study (Booster after Sisonke Study) |
| **Official scientific title** | Phase II randomised open label trial of full and half dose J&J Ad26.CoV2.S and Pfizer BNT162b2 booster vaccinations after receiving the J&J Ad26.CoV2.S prime vaccine through the SISONKE phase IIIB implementation study. |
| **Brief summary describing the background and objectives of the trial** | Open label randomised phase II study of immunogenicity after one of four booster Covid-19 vaccine doses with the J&J Ad26.CoV2.S (full and half dose) and the Pfizer BNT162b2 (full and half dose) vaccine, received after an initial J&J Ad26.CoV2.S prime via the SISONKE phase IIIB implementation study. Participants will be enrolled from the SISONKE phase IIIB implementation study, where a single dose of a J&J Ad26.CoV2.S vaccine was administered to health care workers in South Africa. Health Care Workers (HCW) will be eligible for enrolment if they are ≥30 years of age and we will aim to recruit at least 10% of participants ≥ 55 years of age. Participants will have received the prime J&J Ad26.CoV2.S vaccine through the SISONKE IIIB study at least 4 months prior to enrolment in the BaSiS study. Participants who have previously been infected with SARS-CoV-2 (at least 28 days post symptom resolution or post positive test in asymptomatic) and participants with well controlled comorbidities may be enrolled in the study. In total approximately 300 participants will be enrolled, 75 per prime/boost combination, divided into subgroups per prime/boost combination as follows: 1) 200 participants HIV-uninfected 2) 100 participants who are people living with HIV (PLHIV) Per site: approximately 75 participants will be enrolled. Primary Objectives - To evaluate the immunogenicity of a homologous vaccine boost with either a full (5x 1010 vp/ml, 0.25 ml) or a half dose (2.6x 1010 vp/ml, 0.13 ml) J&J Ad26.CoV2.S, or a heterologous boost, with either a full dose (30mcg, 0.3ml) or a half dose (15mcg, 0.15ml) Pfizer BNT162b2 vaccine, following J&J Ad26.CoV2.S vaccine administered through the SISONKE phase IIIB implementation study by comparing antibody and T cell responses before and after boosting. - To evaluate safety and reactogenicity after a half or full dose J&J Ad26.CoV2.S or Pfizer BNT162b2 vaccine booster dose. Secondary objectives - To assess whether leng |
| **Type of trial** | RCT |
| **Acronym (If the trial has an acronym then please provide)** | BaSiS |
| **Disease(s) or condition(s) being studied** | Infections and Infestations |
| **Sub-Disease(s) or condition(s) being studied** | COVID-19 |
| **Purpose of the trial** | Prevention: Vaccines |
| **Anticipated trial start date** | 08/12/2021 |
| **Actual trial start date** | 08/12/2021 |
| **Anticipated date of last follow up** | 30/09/2022 |
| **Actual Last follow-up date** | |
| **Anticipated target sample size (number of participants)** | 300 |
| **Actual target sample size (number of participants)** | |
| **Recruitment status** | Closed to recruitment,follow-up continuing |
| **Publication URL** | Pending |

| **Secondary Ids** | **Issuing authority/Trial register** |
|---|---|

#### STUDY DESIGN

| **Intervention assignment** | **Allocation to intervention** | **If randomised, describe how the allocation sequence was generated** | **Describe how the allocation sequence/code was concealed from the person allocating the participants to the intervention arms** | **Masking** | **If masking / blinding was used** |
|---|---|---|---|---|---|

| | | | | |
|---|---|---|---|---|
| Parallel: different groups receive different interventions at same time during study | Randomised | Simple randomization using a randomization table created by a computer software program | Allocation was determined by the holder of the sequence who is situated off site | Open-label(Masking Not Used) |

| INTERVENTIONS | | | | | | |
|---|---|---|---|---|---|---|
| **Intervention type** | **Intervention name** | **Dose** | **Duration** | **Intervention description** | **Group size** | **Nature of control** |
| Control Group | Ad26COV2.S | Full (5x 1010 vp/ml, 0.25 ml) | Stat doses at the entry visit | Booster vaccinations, both homologous and heterologous with two different doses of each post J and J Ad26COV2.S prime via SISONKE | 75 | Dose Comparison |
| Experimental Group | Ad26COV2.S | half dose (2.6x 1010 vp/ml, 0.13 ml) | stat dose at entry | Booster vaccine | 75 | |
| Control Group | BNT162b2 | 30mcg (0.3ml) | Stat dose at entry | Booster Covid-19 vaccine doses with the J&J Ad26.CoV2.S (full and half dose) and the Pfizer BNT162b2 (full and half dose) vaccine, received after an initial J&J Ad26.CoV2.S prime via the SISONKE | 75 | Dose Comparison |
| Experimental Group | BNT162b2 | 15mcg (0.15ml) | Stat dose at entry | Booster Covid-19 vaccine doses with the J&J Ad26.CoV2.S (full and half dose) and the Pfizer BNT162b2 (full and half dose) vaccine, received after an initial J&J Ad26.CoV2.S prime via the SISONKE | 75 | |

| ELIGIBILITY CRITERIA | | | | | |
|---|---|---|---|---|---|
| **List inclusion criteria** | **List exclusion criteria** | **Age Category** | **Minimum age** | **Maximum age** | **Gender** |
| 1. Health Care Workers 's who received the J&J Ad26.CoV2.S vaccine through the SISONKE phase IIIB implementation study. 2. Age ≥ 30 years. 3. HIV-uninfected or persons living with HIV who are medically stable on the day of enrolment. 4. Female participants of childbearing potential must have a negative urine BHCG test at screening/enrolment. 5. Willing and able to sign informed consent. 6. Able in the investigator's opinion to comply with study procedures. 7. Plan to stay within reasonable distance of the study site to attend study visits. 8. Participants with well controlled comorbidities, ≤ grade 1 severity at enrolment, including hypertension, type 1 or 2 diabetes, asthma or chronic pulmonary disease, tuberculosis in preceding 3 years, renal disease, cardiac conditions, or autoimmune conditions. | 1. Prior history of thrombotic events including previous deep vein thrombosis. 2. Receipt of any oral or other systemic steroid therapy within the preceding 28 days prior to enrolment. 3. Receipt of any other systemic immunosuppressive agent in the preceding 60 days prior to enrolment. 4. Receipt of any blood transfusion or any blood products within the preceding 6 months prior to enrolment. 5. Receipt of any vaccines in the 28 days prior to enrolment. 6. Cancer either not in remission, or not in remission in preceding 5 years prior to enrolment. 7. Any history of allergic disease or reactions likely to be exacerbated by any component of the vaccine. 8. Any history of hereditary angioedema or idiopathic angioedema. 9. Any history of anaphylaxis secondary to vaccination. 10. Current pregnancy at enrolment. 11. Receipt of any COVID-19 vaccine except for the J&J Ad26.CoV2.S vaccine through the SISONKE phase IIIB implementation study. 12. Symptoms of SARS-CoV-2 disease at the screening /enrolment visit will result in exclusion. 13. Participants not willing to be tested for HIV at entry, unless known to be PLHIV, will be excluded. 14. Enrolment in any other investigational | 80 and over: 80+ Year,Adult: 19 Year(s)-105 Year(s),Aged: 65+ Year(s),Middle Aged: 45 Year(s)-64 Year(s) | 30 Year(s) | 100 Year(s) | Both |

| | |
|---|---|
| | studies for duration of the BaSiS study. |

#### APPROVALS

| Has the study received appropriate ethics committee approval | Date the study will be submitted for approval | Date of approval | Name of the ethics committee |
|---|---|---|---|
| Yes | | 02/11/2021 | Wits Human Research Ethics Committee |

**Ethics Committee Address**

| Street address | City | Postal code | Country |
|---|---|---|---|
| Research Office, Senate House, University of the Witwatersrand, 1 Jan Smuts Avenue, Braamfontein | Johannesburg | 2000 | South Africa |

| Has the study received appropriate ethics committee approval | Date the study will be submitted for approval | Date of approval | Name of the ethics committee |
|---|---|---|---|
| Yes | | 24/11/2021 | SAHPRA |

**Ethics Committee Address**

| Street address | City | Postal code | Country |
|---|---|---|---|
| Building A, Loftus Park, 402 Kirkness Street, Arcadia, | Pretoria | 0001 | South Africa |

| Has the study received appropriate ethics committee approval | Date the study will be submitted for approval | Date of approval | Name of the ethics committee |
|---|---|---|---|
| Yes | | 03/12/2021 | Biomedical Research Ethics Committee UKZN |

**Ethics Committee Address**

| Street address | City | Postal code | Country |
|---|---|---|---|
| Biomedical Research Ethics Committee, Room N40, Govan Mbeki Building, University of KwaZulu-Natal University Road, Westville Campus | Durban | 4000 | South Africa |

#### OUTCOMES

| Type of outcome | Outcome | Timepoint(s) at which outcome measured |
|---|---|---|
| Primary Outcome | - To evaluate the immunogenicity of a homologous vaccine boost with either a full (5x 1010 vp/ml, 0.25 ml) or a half dose (2.6x 1010 vp/ml, 0.13 ml) J&J Ad26.CoV2.S, or a heterologous boost, with either a full dose (30mcg, 0.3ml) or a half dose (15mcg, 0.15ml) Pfizer BNT162b2 vaccine, following J&J Ad26.CoV2.S vaccine administered through the SISONKE phase IIIB implementation study by comparing antibody and T cell responses before and after boosting. - To evaluate safety and reactogenicity after a half or full dose J&J Ad26.CoV2.S or Pfizer BNT162b2 vaccine booster dose. | 2 weeks , 3 months and 6 months |
| Secondary Outcome | - To assess whether length of time between prime and booster dose impacts immunogenicity. - To assess differences in immunogenicity by age and by HIV status. - To evaluate boosted antibody responses against ancestral and novel SARS-CoV-2 strains including D614G, beta, delta, and other variants of concern (VOCs) compared to baseline. - To evaluate the capacity of boosted T cell responses against ancestral and novel SARS-CoV-2 strains including D614G, beta, delta, and other relevant VOCs as they emerge compared to baseline. | 2 weeks, 3 months and 6 months |
| Secondary Outcome | - To evaluate whether clotting profiles in participants at baseline and 2 weeks differ by booster arm, HIV status and age. | 2 weeks , 3 months and 6 months |

#### RECRUITMENT CENTRES

| Name of recruitment centre | Street address | City | Postal code | Country |
|---|---|---|---|---|
| Wits RHI Shandukani CRS | 7 Esselen Street, Hillbrow | Johannesburg | 2001 | South Africa |
| Perinatal HIV Research Unit | Perinatal HIV Research Unit, Soweto Kliptown, Office 7, Walter Sisulu Square, Corner Klipsruit Valley and Union Roads, Kliptown, Soweto, Gauteng | Johannesburg | 1809 | South Africa |

| | | | | |
|---|---|---|---|---|
| CAPRISA eThekwini Clinical Research Site | CAPRISA eThekwini Clinical Research Site, No. 3 Richards Road, Warwick Avenue, Berea, Durban, 4001, KwaZulu-Natal, South Africa | Duban | 4001 | South Africa |
| Masiphumelele Research Site Desmond Tutu Health Foundation | Guinea Fowl Rd, Sunnydale, Fish Hoek, Cape Town, South Africa | Cape Town | 7975 | South Africa |

#### FUNDING SOURCES

| Name of source | Street address | City | Postal code | Country |
|---|---|---|---|---|
| SAMRC | Francie van Zijl Drive, Parowvallei | Cape Town | 7500 | South Africa |

#### SPONSORS

| Sponsor level | Name | Street address | City | Postal code | Country | Nature of sponsor |
|---|---|---|---|---|---|---|
| Primary Sponsor | SAMRC | Francie van Zijl Drive, Parowvallei | Cape Town | 7505 | South Africa | Funding Agency |

#### COLLABORATORS

| Name | Street address | City | Postal code | Country |
|---|---|---|---|---|
| Assoc. Professor Wendy Burgers | 1 Anzio Road, Observatory, 7925, Cape Town | Cape Town | 7925 | South Africa |
| Professor Helen Rees | Cnr Esselen and Klein Street, Hillbrow | Johannesburg | 2001 | South Africa |

#### CONTACT PEOPLE

| Role | Name | Email | Phone | Street address |
|---|---|---|---|---|
| **Principal Investigator** | Lee Fairlie | | +27827809997 | 7 Esselen Street, Hillbrow |
| **City** | **Postal code** | **Country** | **Position/Affiliation** | |
| Johannesburg | 2001 | South Africa | Director of Maternal and Child Health Shandukani Research Centre | |
| **Role** | **Name** | **Email** | **Phone** | **Street address** |
| **Public Enquiries** | Hermien Gous | | +27832833860 | 7 Esselen Street, Hillbrow |
| **City** | **Postal code** | **Country** | **Position/Affiliation** | |
| Johannesburg | 2001 | South Africa | Programme manager | |
| **Role** | **Name** | **Email** | **Phone** | **Street address** |
| **Scientific Enquiries** | Faeezah Patel | | +27835321814 | 7 Esselen Street, Hillbrow |
| **City** | **Postal code** | **Country** | **Position/Affiliation** | |
| Johannesburg | 2001 | South Africa | Site Principal Investigator | |

#### REPORTING

| Share IPD | Description | Additional Document Types | Sharing Time Frame | Key Access Criteria |
|---|---|---|---|---|
| Yes | The results will be shared with participants, policy-makers, relevant stake-holders and will be published in journals and on other public forums. Detailed deidentifies data may be shared with policy-holders specifically if potential changes to COVID vaccine guidelines are indicated. | Clinical Study Report,Informed Consent Form,Statistical Analysis Plan,Study Protocol | Immediately following publication until 12 months post publication | Investigators who apply to review data and are approved by an independent panel |
| **URL** | **Results Available** | **Results Summary** | **Result Posting Date** | **First Journal Publication Date** |
| Not yet available | No | | | |
| **Result Upload 1:** | **Result Upload 2:** | **Result Upload 3:** | **Result Upload 4:** | **Result Upload 5:** |
| **Result URL Hyperlinks** | **Link To Protocol** | | | |
| Result URL Hyperlinks | | | | |

#### Changes to trial information

| Section Name | Field Name | Date | Reason | Old Value | Updated Value |
|---|---|---|---|---|---|
| Trial Information | Recruitment status | 23/08/2022 | End of recruitment period reached. Follow up is ongoing. | Recruiting | Closed to recruitment,follow-up continuing |
| **Section Name** | **Field Name** | **Date** | **Reason** | **Old Value** | **Updated Value** |
| Ethics | Ethics List | 18/01/2022 | SANCTR email requested a re- submission of the document. | True, SAHPRA, Building A, Loftus Park, 402 Kirkness Street, Arcadia,, Pretoria, 0001, South Africa, , 24 Nov 2021, +27125010300,, 5737_10841_4737.pdf, 20210423 | True, SAHPRA, Building A, Loftus Park, 402 Kirkness Street, Arcadia,, Pretoria, 0001, South Africa, , 24 Nov 2021, +27125010300,, 5737_10841_4737.pdf, 20210423 |
| **Section Name** | **Field Name** | **Date** | **Reason** | **Old Value** | **Updated Value** |
| Funding Source | FundingSources List | 18/01/2022 | Updated - did not save at intial submission | | SAMRC, Francie van Zijl Drive, Parowvallei, Cape Town , 7500, South Africa, Government Body, , R29 million, Primary Funder |
