## Supplementary material for "Clot Twist – D-dimer analysis of healthy adults receiving heterologous or homologous booster COVID-19 vaccine after a single prime dose of Ad26.COV2.S in a phase II randomised open-label trial, BaSiS": Basis Protocol v2.0

| | |
|---|---|
| **Trial Title:** | Phase II randomised open label trial of full and half dose J&J Ad26.CoV2.S and Pfizer BNT162b2 booster vaccinations after receiving the J&J Ad26.CoV2.S prime vaccine through the SISONKE phase IIIB implementation study or through the South African COVID-19 vaccination programme. |
| **Study Reference:** | J&J Ad26.COV2.S/Pfizer BNT162b2 (mRNA) prime-boost study<br><br>BaSiS study (**B**ooster **a**fter **Si**sonke **S**tudy) |
| **Protocol Version:** | Version 2.0 |
| **Date:** | 23 March 2022 |
| **Protocol Chair**:<br>**Co-Chairs**: | Lee Fairlie (Wits RHI)<br>Penny Moore (NICD and University of the Witwatersrand)<br>Alex Sigal (AHRI) |
| **Sponsor:** | South African Medical Research Council |
| **Funder:** | South African Medical Research Council |

### Table of Contents


1. Study Team 4
2. Abbreviations 6
3. Synopsis 8
4. Background and rationale 14
    4.1. Background 14
        4.1.1. J&J Ad26.COV2.S 14
        4.1.2. Pfizer BNT162b2 vaccine 16
        4.1.3. Immune response in people living with HIV and other comorbidities 16
        4.1.4. Homologous and Heterologous boost studies 17
        4.1.5. Fractional dosing of vaccines 18
    4.2. Rationale 19
5. Objectives and endpoints 20
    5.1. Primary Objectives 20
    5.2. Secondary objectives 20
    5.3. Exploratory objective 20
    5.4. Endpoint measures 22
6. Methodology 23
    6.1. Trial design 23
    6.2. Study groups 23
    6.3. Trial participants 24
    6.4. Potential risks for participants 24
        6.4.1. Venipuncture 24
        6.4.2. Vaccine reactions 24
        6.4.3. Response to booster 25
        6.4.4. Anaphylactic reactions 25
        6.4.5. Loss of privacy or confidentiality 25
    6.5. Known potential benefits 25
    6.6. Recruitment and Withdrawal of trial participants 26
        6.6.1. Identification of trial participants 26
        6.6.2. Informed consent process 26
        6.6.3. Inclusion and exclusion criteria 27
        6.6.4. Female participants of childbearing potential 28
    6.7. Trial procedures 28
        6.7.1. Observations 28
        6.7.2. Spike binding antibody assays 28
        6.7.3. Neutralisation Assays 29
        6.7.4. T cell assays 29
        6.7.5. Study visits 30
7. Investigational Product 34
    7.1. J&J Ad26.COV2.S adenovector vaccine 34
    7.2. Pfizer BioNTech BNT162b2 modRNA vaccine 34
    7.3. IP preparation, handling, storage and accountability 35
        7.3.1. J&J Ad26.COV2.S adenovector vaccine 35
        7.3.2. Pfizer BioNTech BNT162b2 modRNA vaccine 35
        7.3.3. IP destruction 35
8. Assessment of safety 36
    8.1. Definitions 36
        8.1.1. Adverse Event (AE) 36

8.1.2. Adverse reaction (AR) ... 36
8.1.3. Serious Adverse event (SAE) ... 36
8.1.4. Important medically attended event ... 36
8.1.5. Suspected Unexpected Severe Adverse Reaction (SUSAR) ... 37
8.1.6. Adverse Event of Special Interest (AESI) ... 37
8.2. Grading of events ... 37
8.2.1. Reactogenicity ... 37
8.2.2. Vital signs ... 38
8.2.3. Causality ... 39
8.3. Reporting of AEs ... 39
9. Data Management and Analysis 40
9.1. Randomisation Procedure ... 40
9.2. Sample size ... 40
9.3. Database development ... 41
9.4. Data Entry and Management ... 41
9.4.1. Study-related Forms ... 41
9.4.2. Data Entry and Processing ... 42
9.4.3. Data Security ... 42
9.4.4. Data Quality Control ... 43
9.4.5. Data Transfer ... 43
9.5. Data Analysis ... 44
9.5.1. Immunogenicity analyses ... 44
9.5.2. Safety and Reactogenicity analyses ... 45
10. References 46

### 1. Study Team

Table 1: Protocol team roster

| | |
|---|---|
| South African PI and co-chair | Assoc. Professor Lee Fairlie<br>Wits RHI Shandukani<br>7 Esselen street<br>Hillbrow, 2001<br>Phone: +27827809997<br> |
| Co-chairs | Assoc. Professor Penny Moore<br>HIV Virology Section, National Institute for Communicable Diseases, 1 Modderfontein Road, Johannesburg, South Africa<br>Phone: +27 72 72 44501<br> |
| | Assoc. Professor Alex Sigal<br>Africa Health Institute and University of KwaZulu-Natal<br>719 Umbilo Road, Durban, South Africa<br>Phone: +27 83 446 3092<br> |
| Sponsor | Professor Glenda Gray<br>SA MRC<br>Department of Health, South Africa |
| Funder | SAMRC |
| Trial sites | Wits RHI Shandukani Research Centre<br>Premises 1: 2nd Floor, Hillbrow Health Precinct, Corner Esselen Street and Klein Street, Hillbrow Johannesburg, South Africa, Gauteng, 2001<br>Phone: +27 11 358 5502 Fax: +27 86 548 4889<br><br>Site PI: Dr Faeezah Patel: |
| | PHRU Kliptown<br>Top floor, Walter Sisulu Square, Corner Union and Klipspruit Valley Roads, Kliptown, Soweto, 1809<br>Tel: 011 342 4075<br><br>Site Co-PIs:<br>Clinical Co-PI: Dr Erica Lazarus<br>Non-clinical Co-PI: Anusha Nana |
| | CAPRISA eThekwini Clinical Research Site<br>No. 3 Richards Road, Warwick Avenue, Berea, Durban, 4001, KwaZulu-Natal, South Africa<br>Phone: +27 655 0613<br><br>Site PI: Dr Nigel Garrett |
| | Desmond Tutu Health Foundation: Masiphumelele CRS<br>3 Guinea Fowl Road<br>Sunnydale, Cape Town<br>7975<br>Tel: 021 7853121<br><br>Site PI: Dr Katherine Gill |
| Collaborators | Assoc. Professor Wendy Burgers |

| | |
|---|---|
| | Room 3.36.1, Level 3, Falmouth Building, Faculty of Health Sciences;<br>University of Cape Town, 1 Anzio Road, Observatory, 7925, Cape Town, South Africa<br>Phone: +27825109071<br> |
| **Co-Investigator** | Professor Helen Rees<br>Executive Director, Wits RHI<br>Corner Esselen and Klein Street<br>Hillbrow, 2001<br>Phone: 011 358 5300<br> |

#### 2. Abbreviations

ACLS - Advanced Cardiovascular Life Support

AE - Adverse Event

AR - Adverse Reaction

ART - Antiretroviral Treatment

AESI - Adverse Event of Special Interest

BCEPS – Biometric Co-enrolment Prevention System

BHCG - Beta Human Chorionic Gonadotropin

CAPRISA - Centre for the AIDS Programme in South Africa

ChAd - Chimpanzee Adenovirus

COVID-19 - Coronavirus Disease 2019

FDA - Food and Drug Administration

HCW - Health care worker

HIV - Human Immunodeficiency Virus

ICF - Informed consent form

IPV - Inactivated Polio Vaccine

LICs – Low-income countries

LVNA - live virus neutralisation assay

mAbs - monoclonal antibodies

mRNA - messenger Ribonucleic Acid

nAbs- Neutralising Antibodies

NICD - National Institute for Communicable Diseases

PBMC - Peripheral Blood Mononuclear Cells

PBS - Phosphate-Buffered Saline

PCR - Polymerase Chain Reaction

PHRU - Perinatal HIV Research Unit

PLHIV - People Living with HIV

PNA - Pseudovirus Neutralisation Assay

POC - Point of care

SAE - Serious Adverse Event

SMS - Short Message Service

SUSAR - Suspected Unexpected Serious Adverse Reaction

TMB - Tetramethylbenzidine

UK - United Kingdom

VOC - Variant of concern

VL - Viral Load

Wits RHI - Wits Reproductive Health and HIV Institute

WOCP - Women of childbearing potential

#### 3. Synopsis

| | |
|---|---|
| Title | Phase II randomised open label trial of full and half dose J&J Ad26.COV2.S and Pfizer BNT162b2 booster vaccinations after receiving the J&J Ad26.COV2.S prime vaccine through the SISONKE phase IIIB implementation study. |
| Short title | J&J Ad26.COV2.S**/**Pfizer BNT162b2 prime-boost study |
| Study name | BaSiS study (**B**ooster **a**fter **Si**sonke **S**tudy) |
| Sponsor | South African Medical Research Council (SA MRC)<br>South African Department of Health (DoH) |
| Funder | SA MRC |
| Trial design | Open label randomised phase II study of immunogenicity after one of four booster Covid-19 vaccine doses with the J&J Ad26.COV2.S (full or half dose) or the Pfizer BNT162b2 (full or half dose) vaccine, received after an initial J&J Ad26.COV2.S prime via the SISONKE phase IIIB implementation study or through the South African COVID-19 vaccination programme. |
| Trial participants | Participants who received a single dose of a J&J Ad26.COV2.S vaccine will be recruited from the SISONKE phase IIIB implementation study and through the South African vaccination programme. . Participants will be eligible for enrolment if they are 18 years of age and we will aim to recruit at least 10% of participants ≥ 55 years of age. Participants will have received the prime J&J Ad26.COV2.S vaccine at least 4 months prior to enrolment in the BaSiS study. Participants who have previously been infected with SARS-CoV-2 (at least 28 days post symptom resolution or post positive test in asymptomatic) and participants with well controlled comorbidities may be enrolled in the study. In total approximately 300 participants will be enrolled, 75 per prime/boost combination, divided into subgroups per prime/boost combination as follows:<br>1) Approximately 200 participants HIV-uninfected<br>2) At least 100 participants who are people living with HIV (PLHIV)<br>Per site: approximately 75 participants will be enrolled, a site may enrol up to 150 participants. |
| Sample size | 300 participants, approximately 75 at each site will be enrolled, however each site may enrol up to 150 participants.<br>**Group A** = 75 Ad26.COV2.S prime plus full dose Ad26.COV2.S booster (50 HIV- 25 HIV+) at a dose of 5x $10^{10}$ vp/ ml (0.25 ml)<br>**Group B** = 75 Ad26.COV2.S prime plus half dose Ad26.COV2.S booster (50 HIV- 25 HIV+) at a dose of 2.6x $10^{10}$ vp/ ml (0.13 ml)<br>**Group C** = 75 Ad26.COV2.S prime plus full dose BNT162b2 booster (50 HIV-, 25 HIV+) at a dose of 30mcg (0.3 ml)<br>**Group D** = 75 Ad26.COV2.S prime plus half dose BNT162b2 booster (50 HIV-, 25 HIV+) at a dose of 15mcg (0.15 ml) |
| Planned trial period | Study enrolment will be conducted over 4 – 6 months. Follow-up visits will take place 2 weeks, 3 months, and 6 months (study exit) from the enrolment date. Interim visits may be arranged for any solicited adverse events (AEs) beyond 7 days post vaccination, or unsolicited AEs throughout the study post vaccination if they are grade 3 or higher. |

| | |
|---|---|
| | Additional study visits will be conducted for participants who develop symptoms of SARS-CoV-2 infection or test positive for SARS-CoV-2 infection with a nasopharyngeal swab polymerase chain reaction test or rapid antigen test while on the BaSiS study.<br>An interim analysis for immunogenicity will be performed after the 3-month visit. The highest GMT of neutralising antibodies, calculated per arm for each of the four arms, will be used as the benchmark for comparison to individual immunogenicity responses. The GMT of neutralising antibodies from the 3-month post-vaccination visit will be utilised for this purpose. If insufficient immunogenicity (defined as <75% of the geometric mean titre (GMT)) is observed between the full dose and half dose arms, or between J&J Ad26.COV2.S and Pfizer BNT162b2 arms, regardless of dose, the homologous or heterologous booster vaccine eliciting the most robust response will be offered at the 6-month visit. Any participant, in any study arm (including the arm that elicits the highest GMT), will be offered the booster that elicited the highest GMT, if their 3-month neutralising antibody level is <75% of the highest GMC. Participants will be contacted by the study staff and will be offered a booster dose if these criteria are met. The study Data and Safety Monitoring Board will review data and adjudicate on this process. |

**Objectives and endpoint measures**

*Primary Objectives*

- To evaluate the immunogenicity of a homologous vaccine boost with either a full (5x $10^{10}$ vp/ml, 0.25 ml) or a half dose (2.6x $10^{10}$ vp/ml, 0.13 ml) J&J Ad26.COV2.S, or a heterologous boost, with either a full dose (30mcg, 0.3ml) or a half dose (15mcg, 0.15 ml) Pfizer BNT162b2 vaccine, following J&J Ad26.COV2.S vaccine administered through the SISONKE phase IIIB implementation study or SA vaccination programme, by comparing antibody and T cell responses before and after boosting.
- To evaluate safety and reactogenicity after a half or full dose J&J Ad26.COV2.S or Pfizer BNT162b2 vaccine booster dose.

*Secondary objectives*

- To assess whether length of time between prime and booster dose impacts immunogenicity.
- To assess differences in immunogenicity by age and by HIV status.
- To evaluate boosted antibody responses against ancestral and novel SARS-CoV-2 strains including D614G, beta, delta, and other variants of concern (VOCs) compared to baseline.
- To evaluate the capacity of boosted T cell responses against ancestral and novel SARS-CoV-2 strains including D614G, beta, delta, and other relevant VOCs as they emerge compared to baseline.

*Exploratory objective*

- To evaluate whether clotting profiles in participants at baseline and 2 weeks differ by booster arm, HIV status and age.

Table 2: Objectives and endpoint measures for the BaSiS study

| | Objectives | Endpoint measures |
|---|---|---|
| **Primary** | Immunogenicity (antibody and T cell) | Neutralisation titres post-boost versus baseline using the pseudovirus neutralisation and live virus neutralisation assays. T cell response magnitudes post-boost versus baseline using intracellular cytokine staining. |
| | Safety and reactogenicity | Local and systemic reactogenicity measured by diary cards.<br>Monitor ARs, SAEs, SUSARS, AESI |
| **Secondary** | Evaluate impact of duration from prime to boost on immunogenicity | Antibody and T cell immunogenicity in each group from time of prime to boost |
| | Differences in immunogenicity by HIV status and age | Comparison of antibody and T-cell response between PLHIV and HIV-uninfected and those ≥ 55 years and < 55 years |
| | Evaluate cross-reactivity of boosted neutralising antibody responses to ancestral and novel SARS-CoV-2 variants compared to baseline | Pseudovirus and live virus neutralising antibody titres pre and post booster vaccine, tested against D614G, delta, beta (and other VOCs should these become locally relevant) |
| | Evaluate cross-reactivity of T cell responses to ancestral and novel SARS-CoV-2 variants compared to baseline | T cell responses to D614G, delta, beta (and other VOCs should these become locally relevant) |
| **Exploratory** | Evaluate differences in clotting profiles with different boosters, age and HIV status | Compare differences between haemoglobin, platelets and D-dimer at baseline and 2 weeks in each study arm and according to HIV status and age |

Schedule of evaluations for the BASIS study

Table 3: Schedule of evaluations for the BaSiS study

| | **Screening/ enrolment (Day 0)** | **2 weeks** | **3 months** | **6 months** | **Interim visit [a]** | **Illness visit** |
|---|---|---|---|---|---|---|
| Visit window | NA | ±3 days | ±7 days | ±14 days | N/A | |
| ICF | X | | | | | |
| Weight, height, body mass index | X | | | | | |
| Vital signs [b] | X | X | X | X | X | X |
| Medical history | X | X | X | X | X | X |
| COVID-19 infection related history | X | X | X | X | X | X |
| Targeted physical examination if necessary | X | X | X | X | X | X |
| Contraception history, LMP and Urine BHCG in WOCP | X | | | | | |
| Eligibility check | X | | | | | |
| Vaccination check [c] | X | X | X | X | X | |
| HIV Elisa test [d] | X | | | | | |
| FBC and D-Dimers | X | X | | | X | |
| Immunology bloods in PLHIV (CD4, CD8, lymphocytes, neutrophils) | X | | | X | | |
| HIV-1 Viral Load in PLHIV | X[e] | X[f] | | X | | |
| Immunogenicity bloods-SARS-CoV-2 antibodies [g] | X | X | X | X | | |
| Immunogenicity bloods-Cellular immunity (PBMC) [g] | X | X | X | X | | |
| Nasal swabs SARS-CoV-2 PCR | X | | | | | X |
| Randomisation | X | | | | | |
| Vaccination | X | | | | | |
| Post-vaccination observations (15-30 mins) | X | | | | | |
| Train on and issue vaccine diary card | X | | | | | |
| Train on & issue memory aid | X | | | | | |
| Diary card collection | | X | X | | | |
| Illness diary issue | | | | | | X |
| Collect solicited and unsolicited AEs | | X | X | X | X | X |
| Blood volumes | PLHIV: 70ml<br>HIV-: 60 ml | 60 ml<br>(+ 6 ml for newly diagnosed PLHIV) | 50 ml | PLHIV: 70 ml<br>HIV-: 60 ml | | |

[a] Interim visits will be arranged in the following circumstances:

- Reactogenicity event greater or equal to grade 3 or any reactogenicity event ongoing at day 7.
- Solicited AEs beyond 7 days post vaccination, or unsolicited AEs throughout the study post vaccination if they are grade 3 or higher
- Clotting abnormalities grade 2 or higher at baseline and week 2 or symptomatic between vaccination and the 2 week visit

[b] Include blood pressure, pulse rate, saturations on room air, respiratory rate, temperature (infrared tympanic or oral with a plastic sleeve cover, to minimise infection risk).
[c] Brief eligibility check to evaluate whether any additional COVID-19 vaccines or any other vaccines received during study period.
[d] Counselling and referral for HIV care including antiretroviral therapy if a participant tests positive on HIV Elisa.
[e] VL will be done at enrolment in people who are known to be living with HIV
[f] VL will be done in people who are diagnosed with HIV at enrolment visit at the 2-week visit
[g] Elisa testing will be done on all samples, neutralisation assays will only be done on participants who test positive on Elisa.

### 4. Background and rationale

#### 4.1. Background

In March 2020 a global pandemic of COVID-19 caused by the SARS-CoV-2 virus was declared, and subsequently over 200 million people have been confirmed as SARS-CoV-2 positive and over 4 million deaths have been directly attributed to COVID-19 globally, with 2.5 million cases and over 80,000 deaths reported in South Africa (1). The ancestral SARS-CoV-2 strain has mutated over time and variants of concern (VOC) have emerged including the B.1.1.7 (Alpha), B1.351 (Beta), B.1.617.2 (Delta) and P.1 (Gamma) variants (2) and more recently the B.1.1.529 BA.1 and BA.2 (Omicron) variants identified in November 2021 in South Africa (3). In South Africa, the beta variant was predominant in the second wave of infection, and the delta variant was dominant in the third wave, with both variants known to have increased transmissibility and escape from neutralisation by convalescent and vaccine induced antibody immunity. The delta variant appears to result in more severe disease relative to the alpha variant, with data from Scotland reporting a risk of hospitalisation with delta variant infection double that of the alpha variant (4). In pregnant women, alpha and delta variant strains result in increased severity of maternal disease and adverse pregnancy outcomes compared to the ancestral strain (5). The omicron variant, largely responsible for the fourth wave, displayed high transmissibility but apparently milder disease with lower rates of severe disease, hospitalisation and death (6, 7). However, Omicron-triggered neutralisation in unvaccinated SARS-CoV-2 naïve individuals is not extensively cross-reactive to VOCs, leaving these individuals likely to be more susceptible to reinfection by circulating and emerging VOCs that may potentially be more virulent(8),. Throughout 2020, tremendous efforts to develop effective, safe COVID-19 vaccines resulted in emergency use authorization of vaccines with different mechanisms of action, and more recently licensure of some vaccines. Among these are the single dose Johnson & Johnson (J&J) adenoviral vectored J&J Ad26.COV2.S vaccine and the two-dose Pfizer BNT162b2 mRNA vaccines, both of which use the SARS-CoV-2 ancestral spike glycoprotein sequence as the antigen. Other vaccine types include inactivated virus vaccines (Vaxzevria/AstraZeneca/ChAdOx, CoronaVac, Covaxin and others), and protein subunit vaccines such as the Novavax NVX-CoV2373 (9). Globally, over 5 billion vaccine doses have been administered, almost 30% of the world's population have received at least one vaccine dose and over 14% have completed vaccination. However, in low-income countries, less than 10% of people have been vaccinated (10), resulting in a need for strategies to accelerate vaccine coverage in the context of limited vaccine supplies and the high cost of vaccines. South Africa has expanded vaccine access in a stepwise manner: initially single dose J&J Ad26.COV2.S to health care workers then those with high exposure such as schoolteachers and the police force, followed by either the two-dose Pfizer BNT162b2 vaccine regimen or single dose J&J Ad26.COV2.S vaccine to those over 60 years of age, then to over 35-year-olds, then to all people over 18-years of age (from 1st of September 2021). In October 2021, the age of eligibility for Pfizer BNT162b2 was expanded to include 12-18 year olds. Since booster doses were introduced late December 2021 and since February 2022, homologous and heterologous booster vaccines have been made available for those previously vaccinated. Here we focus on the J&J Ad26.COV2.S and Pfizer BNT162b2, currently approved in South Africa.

##### 4.1.1. J&J Ad26.COV2.S

The J&J Ad26.COV2.S vaccine produced by Johnson & Johnson is a single dose, replication-incompetent, recombinant, adenovirus serotype 26 vectored SARS-CoV-2 spike protein vaccine. The safety and efficacy of the J&J Ad26.CoV.S was assessed globally in the ENSEMBLE phase 3 trial, a randomised, placebo-controlled, double-blind study (11). The study enrolled 44 000 participants, almost 7000 from

South Africa, and demonstrated an overall vaccine efficacy against moderate to severe disease of 66.9% (adjusted 95% confidence interval [CI], 59.0 to 73.4) (11). The vaccine had greater efficacy against severe and critical COVID-19 for those infected ≥14 days post vaccine, 76.7% (adjusted 95% CI, 54.6 to 89.1), for those with onset of disease ≥28 days post vaccine, 85.4% efficacy (adjusted 95% CI, 54.2 to 96.9)(11). In South Africa, where during the Ensemble trial the majority (94.5%) of SARS-CoV-2 infections were due to the beta variant (20H/501Y.V2), vaccine efficacy against infections ≥14 days after the dose was 52.0% and ≥28 days, 64.0%, against moderate-to-severe/critical SARS-CoV-2 infection. Against severe-critical infection, vaccine efficacy was 73.1% at ≥14 days and 81.7% at ≥28 days(11). As expected, reactogenicity was higher in the vaccine compared to placebo group but most events were mild to moderate and serious adverse events were distributed across arms, with no SARS-CoV-2 related deaths in the vaccine arm (11).

Following on from the J&J ENSEMBLE study, in South Africa approximately half a million health care workers received the single dose J&J Ad26.COV2.S vaccine through the SISONKE phase IIIB implementation study (12). Interim data from the study showed only around 2% of vaccinees reported any adverse events and that thrombocytopenic thromboembolism, initially reported with the ChAdOx1 nCoV-19 vaccination, also adenovirus based, was extremely rare and occurred in only 1.7/100 000 participants, (13). Recent data showed approximately 2% of HCW had breakthrough infection with SARS-CoV-2, mostly mild (95.6%), with 2.76% moderate, 0.49% severe and 0.4% resulting in death; 76% of people who died had comorbidities (12). The single J&J Ad26.COV2.S vaccine resulted in 65-66% protection against hospitalisation and 91-96% protection against death (12). These results were reported over the time of the beta and delta variants, suggesting a good response across both variants. In addition, at 8 months post vaccine, Barouch et al., report sustained durability and neutralising antibody breadth expansion over this time period, suggestion B-cell response maturation without additional boosting (14). Data from the J&J Ad26.COV2.S phase 1/2a study (COV1001) similarly show that neutralising antibodies remain stable until 8-9 months post prime, and binding antibodies until 6 months post prime, regardless of age (15). Additional doses of the J&J Ad26.COV2.S vaccine have been made available to South Africa and other sub-Saharan African countries.

Although the J&J Ad26.COV2.S vaccine as a single dose is compelling programmatically due to reduced costs, simpler storage requirements and less complex logistics, especially for health systems in resource constrained settings, the prevalence of breakthrough infections, although largely mild, indicated that protection against infection might need to be improved in Ad26.COV2.S vaccinees. In addition, maintaining protection from severe disease over time, in light of the emergence of VOC, is essential. Protection from infection correlates with high antibody titres elicited by vaccines (16) and reduced neutralisation titres have been reported for J&J Ad26.COV2.S vaccines against the beta, delta and omicron variants compared to the ancestral variant (17). This prompted the authors of these studies to suggest that a second J&J booster or a heterologous prime-boost strategy may enhance titres and reduce infections, and ensure ongoing and durable protection following immunisation (17). SISONKE-2 enrolled 227 310 HCW from the original SISONKE implementation study prior to the start of the Omicron fourth wave in South Africa. Participants were given a homologous boost of the J&J Ad26.COV.2.S vaccine 6-9 months after their initial vaccination. After adjusting for confounders, VE was shown to increase over time with 85% prevention of hospitalisation at 1-2 months post-boost (18). Given the benefits of booster vaccination in the context of inequity of vaccine access in Africa, understanding whether fractional dosing, further described in section 4.1.5, will produce similarly robust immune responses is critical.

##### 4.1.2. Pfizer BNT162b2 vaccine

The Pfizer BNT162b2 vaccine, a messenger RNA (mRNA) vaccine which encodes the secreted trimerised SARS-CoV-2 spike protein, has also had extremely favourable results. In the phase I study, the Pfizer BNT162b1 and Pfizer BNT162b2 vaccines were evaluated at doses of 10mcg, 20mcg, 30mcg and 100mcg. Vaccines were administered 21 days apart, except the 100mcg dose which was a single dose, to evaluate safety and immunogenicity. The Pfizer BNT162b2 vaccine at a 30mcg dose was selected for further development, due to the slight decrease in reactogenicity compared to the Pfizer BNT162b1 vaccine and favourable immunogenic responses for SARS-CoV-2 neutralising antibody, although the 20mcg dose also performed well, with similar responses compared to 30mcg (19). The phase II/III multinational randomised placebo-controlled study enrolled over 43 000 participants and evaluated safety and efficacy, demonstrating a 95% efficacy and favourable safety profile for the Pfizer BNT162b2 vaccine (20). Globally hundreds of millions of people, over 200 million in the United States alone, 18 years and older have received this vaccine (21). Adverse events surveillance has reported anaphylaxis - 11.1 per million cases meeting Brighton Classification case definitions, and myocarditis - 40.7 per million cases, which has been reported more commonly in males less than 30 years of age (22, 23). Benefit risk analyses have been done and found that the risks of COVID-19 disease far outweigh the comparatively small risks of these adverse events (22, 23). A study evaluating the effectiveness of mRNA vaccines in HCWs including the Pfizer BNT162b2 and the Moderna vaccine in the United States, reported a vaccine efficacy of 82% (95%CI = 74%–87%) after a single dose and 94% (95% CI = 87%–97%) after 2 doses (24). “Real world” effectiveness data from national surveillance in Israel in the first four months of the Pfizer BNT162b2 vaccine rollout reported adjusted rates of effectiveness from 7 or more days after the second vaccine (25). They report 95.3% (95% CI 94.9–95.7) effectiveness overall, 91.5% (95% CI 90.7–92.2) effectiveness against asymptomatic SARS-CoV-2 infection; 97.0% (95% CI 96.7–97.2) for symptomatic COVID-19 infection; 97.2% (95% CI 96.8–97.5) for hospitalisation related to COVID-19 and 96.7% (95% CI 96.0–97.3) effectiveness against death related to COVID-19 (25). When evaluating effectiveness against variant strains, Bernal et al., reported a slight decrease in effectiveness in people vaccinated with two doses of the Pfizer BNT162b2 vaccine between the alpha [93.7% (95% CI, 91.6 to 95.3)] and delta variant [88.0% (95% CI, 85.3 to 90.1)] (26). This suggests a slight but insignificant loss in effectiveness with the delta variant, but still high levels of effectiveness against this variant (26). Immunogenicity data supports this high level of efficacy and effectiveness, with the Pfizer BNT162b2 vaccine eliciting extremely high titres against the original variant, and modest preservation of neutralisation activity against VOCs including beta and delta (17). In South Africa, data from the fourth omicron-driven COVID-19 wave showed a 70% (95% CI, 62 to 76) vaccine efficacy against the Pfizer BNT162b2 vaccine for hospitalisation, a reduction from 93% (95% CI, 90 to 94) in previous waves(27). Despite the number of spike protein mutations in the Omicron variant, increasing transmissibility and reduction in neutralising antibodies, T-cell responses to both Pfizer BNT162b2 and J&J Ad26.COV2.S appear maintained and similar to that seen in previous waves (28).

##### 4.1.3. Immune response in people living with HIV and other comorbidities

South Africa has the largest HIV epidemic in the world, with 7.7 million people living with HIV (PLHIV) with an overall prevalence of about 19% and approximately 70% on antiretroviral therapy (ART) (29). Immunosuppression caused by HIV is known to interfere with protective vaccination against multiple pathogens, typically as a consequence of sub-optimal antibody responses (30-33).

In line with this, results from a South-African phase IIb trial of the Novavax NVX-CoV2373 vaccine, which uses a stabilised prefusion spike protein, showed 60% efficacy in HIV-uninfected individuals. However, overall efficacy dropped to 49% upon inclusion of PLHIV, although it is important to note that the numbers of PLHIV in the study were very small (34). Nonetheless, there were more breakthrough cases in PLHIV in the vaccine arm than the placebo arm. A small study evaluating immune response in PLHIV compared to HIV-uninfected controls 7-17 days after the second Pfizer BNT162b2 vaccine showed similar spike protein binding antibody titres, neutralising antibody to the vaccine spike protein as well as variants of concern including the beta and delta variants (35). Cellular immune responses between the groups were similar (35). In South Africa, 104 PLHIV well controlled on ARVs, were enrolled in the ChAdOx1 nCoV-19 study and had comparable tolerability, safety and immunogenicity responses to the vaccine compared to HIV-uninfected controls (36).

These results make it imperative to measure vaccine effectiveness in PLHIV both to prevent damage to the health and wellbeing of PLHIV, and for effective control of the SARS-CoV-2 pandemic in South Africa.

In addition to HIV, older age and other prevalent comorbidities may affect vaccine effectiveness in the South African setting. It is well established that increasing age is associated with weakened immune responses and increased risk of infection, severe disease, and death. Obesity, hypertension, and diabetes mellitus are known risk factors for severe disease and death due to SARS-CoV-2 infection. In the Pfizer BNT162b2 vaccine phase II and III studies, just over 20% of people had comorbidities, most commonly hypertension, diabetes, and chronic lung disease. The vaccine showed similar efficacy in people with and without comorbidities and adverse events were not increased in those with comorbidities (37).

###### 4.1.4. Homologous and Heterologous boost studies

Booster vaccination is required for multiple vaccines including most childhood vaccinations (38). With the exception of the J&J Ad26.CoV2 single dose vaccine, the majority of COVID-19 vaccines are comprised of a homologous two dose primary vaccine schedule, including the ChAdOx adeno-vectored and mRNA vaccines (39). Mouse models have demonstrated an increased immunogenic response with an adeno-vectored prime and mRNA vaccine boost compared to an adeno-vectored boost(40). Recently, compelling data has emerged that a heterologous boost with an mRNA-based vaccine such as the Pfizer BNT162b2 elicits a strong immunogenic response when following the adeno-vectored ChAdOx-1 vaccine prime (39, 41). In a recent phase II randomised trial in Spain, data on 676 participants who received a single ChAdOx-1 vaccine and were randomised to receive a Pfizer BNT162b2 boost was reported. The control group received no booster vaccine. The response ratio of heterologous boost:control arm was 77·69 (95% CI 59·57–101·32) for RBD protein and 36·41 (29·31–45·23) for trimeric spike protein IgG. In this study, participants reported mainly mild (68%) and moderate (30%) reactogenicity symptoms, most commonly injection site pain and induration, myalgia and headache with no serious adverse events reported (41). The CoM-COV study in the UK followed a prime-boost design with eight arms, including randomisation to ChAdOx-1/ ChAdOx-1, ChAdOx-1/Pfizer BNT162b2, Pfizer BNT162b2/ChAdOx-1 and Pfizer BNT162b2/ Pfizer BNT162b2 (39). In 463 participants, the ChAdOx-BNT162b2 regimen boosted anti-spike IgG 9.3-fold compared to those receiving two doses of ChAdOx-1 vaccine. Cellular responses, as measured by IFN-gamma ELISPOT, were also 3.9-fold greater in the heterologous prime-boost regimen (39). These results showed non-inferiority of the heterologous ChAdOx-1/Pfizer BNT162b2 prime-boost strategy, to the homologous ChAdOx-1/ ChAdOx-1 strategy. In the Pfizer BNT162b2/ChAdOx-1 and Pfizer BNT162b2/ Pfizer BNT162b2 arms, heterologous boost did not achieve non-inferiority, but SARS-CoV-2 anti-spike IgG concentrations were higher in the heterologous arm (39).

The rate of systemic adverse events and paracetamol use in the two heterologous arms was higher than the homologous arms with 40 (36%) participants in the ChAdOx-1/ ChAdOx-1, 63/110 (57%) in the ChAdOx-1/Pfizer BNT162b2 arm, 48/117 (41%) in the Pfizer BNT162b2/ Pfizer BNT162b2 arm and 68/114 (60%) in the Pfizer BNT162b2/ChAdOx-1 reporting paracetamol use which mirrored reactogenicity (42). Safety blood tests showed similar results between the four arms (42). More recent data from the US, where 456 participants were enrolled who had received one of three covid-19 prime vaccines (BNT162b2, J&J Ad26.COV2.S or mRNA-1273) received a homologous or heterologous booster vaccine at least 12 weeks after their prime vaccine. In this study, heterologous booster vaccines demonstrated an equivalent or higher binding and neutralising antibody response compared to heterologous, reactogenicity did not differ between the arms and there were no safety concerns(43). Therefore, heterologous prime-boost vaccination has the potential to be more cost effective since it increases vaccine efficacy, uses a lower cost prime, be more available since the adeno-vectored vaccines are more stable and available, and increase the effectiveness of vaccination. This approach warrants further evaluation particularly with the J&J Ad26.COV2.S vaccine as a prime, where data is not currently available.

###### 4.1.5. Fractional dosing of vaccines

Fractional dosing of vaccines has the advantage of reducing the amount of vaccine required. At an individual level, this has the potential to reduce reactogenicity and adverse events following immunisation. Reduced use of vaccines is advantageous in the context of vaccine shortages such as with COVID-19 vaccines and when vaccine prices remain high and unaffordable in many low-income countries (LICs). The cost would be reduced, and the amount of available vaccines can be distributed more widely. This approach was used with the 17D yellow fever vaccine during the 2016 outbreak in Kinshasa where there was a shortage of yellow fever vaccine (44). One fifth fractional dosing of the vaccine was used for children over 2 years of age and non-pregnant adults. Fractional dosing had a good safety profile and at 1 month follow-up, 98% of participants overall were seropositive, and 98% of participants who were seronegative before receiving vaccine had seroconverted (44, 45). Seropositivity was sustained in 97% of the participants who remained in follow-up at 12 months (45). A similar approach is being evaluated for inactivated polio vaccine (IPV). A meta-analysis and systematic review by Mashunye et al., reported no significant difference in seroconversion when two or three doses of fractional IPV delivered intradermally, compared to full dose IPV was given intramuscularly. Full dose IPV dose results in higher poliovirus type 1, 2 and 3 antibody titres, however the authors conclude that fractional IPV dosing is feasible and can reduce the demand on vaccine supply and potentially the associated cost (46).

Given the shortages of availability of COVID-19 vaccines, the associated cost and lack of vaccine equity, particularly in LICs where less than 10% of the population has been vaccinated, fractional dosing particularly for a prime-boost strategy is advantageous. The Pfizer BNT162b2 vaccine has demonstrated a robust immunogenicity response, even at lower doses in phase 1 studies and since the adenovirus vectored-mRNA prime boost strategy has demonstrated good results with a ChAdOx prime and Pfizer BNT162b2 vaccine boost this strategy should be further explored (19, 41). The ongoing Phase 1/2a study (COV1001, NCT04436276) recently reported results after evaluating a full ($5x10^{10}$ viral particles [vp]) and quarter ($1.25x10^{10}$ vp) J&J Ad26.COV2.S boost at 6 months post Ad26.COV2.S prime (15). The full dose booster elicited a 9-fold increase in spike binding protein antibodies at 7 days post boost compared to 29 days post prime, and the quarter dose a 6-7.7 fold increase at day 28 post boost, compared to day 29 post prime (15). The response to quarter dose boost was slower in participants over 65 years, but at 28 days was similar in younger and older participants (15). Solicited local adverse events were similar between the prime and quarter dose booster, while solicited systemic adverse events were lower after the boost, 61.7% vs 28% (15).

This study evaluates a prime-boost strategy in HCW and people vaccinated through the SA vaccination programme in South Africa, who received a J&J Ad26.COV2.S vaccine prime through the SISONKE phase IIIB implementation study and routine programmes, receiving either a full dose Ad26.COV2.S booster ($5x\ 10^{10}$ vp/ml) or a half dose Ad26.COV2.S booster ($2.6x\ 10^{10}$ vp/ml) or a full (30mcg) or half (15mcg) dose Pfizer BNT162b2 booster vaccine. The study includes people over 18 years of age, regardless of HIV status.

#### 4.2. Rationale

1. Ad26.COV2.S prime alone triggers lower titres of neutralising antibodies than mRNA vaccines.
2. Other heterologous combinations of adenovirus vectored prime (ChAdOx) and RNA vaccine boosts (Pfizer BNT162b2) are highly immunogenic.
3. Although data exists for J&J Ad26.COV2.S vaccine prime followed by an mRNA vaccine boost, data including fractional homologous and heterologous dosing remains scarce.
4. There is a scarcity of data regarding the immunogenicity of vaccines in PLHIV, who account for a significant proportion of South Africans.
5. People over the age of 55 years have a progressively weakened immune response to vaccines and may benefit from a homologous or a heterologous prime boost.
6. People living with HIV have compromised immune responses even if well controlled on ARVs and may benefit from a homologous or heterologous prime boost.
7. There is public health benefit, including cost and increased availability for greater numbers of people, in addition to potential reduction in local and systemic adverse effects, with fractional dosing for booster vaccines. Available data, although limited, demonstrates that fractional dosing results in robust immune responses.

To boost the immunogenicity of the J&J Ad26.COV2.S vaccine already administered in the SISONKE phase IIIB implementation study and the SA vaccination programme, we propose to test a J&J Ad26.COV2.S /J&J Ad26.COV2.S homologous prime-boost vaccination design and a J&J Ad26.COV2.S/Pfizer BNT162b2 heterologous prime-boost vaccination design, which will include 4 arms:

A. J&J Ad26.COV2.S/ J&J Ad26.COV2.S full dose homologous prime-boost vaccination
B. J&J Ad26.COV2.S/ J&J Ad26.COV2.S half dose homologous prime-boost vaccination
C. J&J Ad26.COV2.S/Pfizer BNT162b2 full dose heterologous prime-boost vaccination
D. J&J Ad26.COV2.S/Pfizer BNT162b2 half dose heterologous prime-boost vaccination

This design will determine if a boost of J&J Ad26.COV2.S or Pfizer BNT162b2 mRNA vaccine triggers high levels of nAbs and T cells, and determine whether the J&J Ad26.COV2.S and Pfizer BNT162b2 dosage can be reduced for cost effectiveness/vaccine sparing, and reduction in local and systemic adverse events post-vaccination.

Immunogenicity will be quantified by measuring nucleocapsid binding antibodies (to assess prior infection) and neutralisation capacity of blood plasma, a measure highly correlated to vaccine efficacy (16). Trial participants will be HCWs already vaccinated with one dose of J&J Ad26.COV2.S. Trial participants will be enrolled and have blood draws at baseline drawn immediately before vaccination, and then followed up at 2 weeks, 3 months and 6 months post vaccination to test peak immunogenicity and durability. Peripheral blood mononuclear cells will be isolated for evaluating T cell immune responses.

Safety evaluations will be measured with the participant-administered diary card, recording local and systemic adverse reactions to the vaccine. Although one heterologous boost study reports an increase in reactogenicity compared to homologous boost, which may be a short-term disadvantage, these adverse events are likely short-lived and manageable particularly with the administration of agents such as paracetamol(42). It will be important to establish short- and longer-term adverse reactions with a J&J Ad26.COV2.S-Pfizer BNT162b2 prime-boost strategy, given that this information cannot be extrapolated from current clinical trials. Such information will assist with decision-making regarding prime-boost strategies in South African and sub-Saharan African countries where this strategy may be most likely given the currently available vaccines being offered in the rollout.

Binding antibodies will be determined by ELISA to the nucleocapsid. Neutralisation capacity will be determined independently by the laboratory of Penny Moore at NICD using a pseudovirus neutralisation assay (PNA) and Alex Sigal at Africa Health Research Institute using a live virus neutralisation assay (LVNA) by established protocols (47). SARS-CoV-2 variants to be neutralised will include the ancestral reference strain and the beta, delta and omicron variants for all samples, as well as other variants which may emerge for select samples. Assays will be completed once the effect of boost on neutralisation at the tested significance level outlined in the analysis plan is determined. T cell responses to the ancestral, beta, delta, omicron and other VOC will be measured by Wendy Burgers at the University of Cape Town, as described (48). Interim results will be made available after the 2 week and 3 months post-boost timepoints. Study follow up and evaluation will be extended through 6 months to assess durability of the booster immune response.

Trial sites will be Wits RHI Shandukani and PHRU Kliptown in Johannesburg, the CAPRISA eThekwini Clinical Research Site in Durban and the Desmond Tutu Health Foundation site in Masiphumelele in Cape Town.

### 5. Objectives and endpoints

#### 5.1. Primary Objectives

- To evaluate the immunogenicity of a homologous vaccine boost with either a full (5x $10^{10}$ vp/ml, 0.25 ml) or a half dose (2.6x $10^{10}$ vp/ml, 0.13 ml) J&J Ad26.COV2.S, or a heterologous boost, with either a full dose (30mcg, 0.3 ml) or a half dose (15mcg, 0.15 ml) Pfizer BNT162b2 vaccine, following J&J Ad26.COV2.S vaccine administered through the SISONKE phase IIIB implementation study and SA vaccination programme by comparing antibody and T cell responses before and after boosting.
- To evaluate safety and reactogenicity after a half or full dose J&J Ad26.COV2.S or Pfizer BNT162b2 vaccine booster dose.

#### 5.2. Secondary objectives

- To assess whether length of time between prime and booster dose impacts immunogenicity.
- To assess differences in immunogenicity by age and by HIV status.
- To evaluate boosted antibody responses against ancestral and novel SARS-CoV-2 strains including D614G, beta, delta, and other variants of concern (VOCs) compared to baseline.
- To evaluate the capacity of boosted T cell responses against ancestral and novel SARS-CoV-2 strains including D614G, beta, delta, and other relevant VOCs as they emerge compared to baseline.

#### 5.3. Exploratory objective

- To evaluate whether clotting profiles in participants at baseline and 2 weeks differ by booster arm, HIV status and age.

#### 5.4. Endpoint measures

Table 4: Objectives and endpoint measures for the BaSiS study

| | Objectives | Endpoint measures |
|---|---|---|
| **Primary** | Immunogenicity (antibody and T cell) | Nucleocapsid binding antibody titres. Neutralisation titres post-boost versus baseline using the pseudovirus neutralisation and live virus neutralisation assays. T cell response magnitudes post-boost versus baseline using intracellular cytokine staining. |
| | Safety and reactogenicity | Local and systemic reactogenicity measured by diary cards.<br>Monitor ARs, SAEs, SUSARS, AESI |
| **Secondary** | Evaluate impact of duration from prime to boost on immunogenicity | Antibody and T cell immunogenicity in each group from time of prime to boost |
| | Differences in immunogenicity by HIV status and age | Comparison of antibody and T-cell response between PLHIV and HIV-uninfected and those ≥55 years and < 55 years |
| | Evaluate cross-reactivity of boosted neutralising antibody responses to ancestral and novel SARS-CoV-2 variants compared to baseline | Pseudovirus and live virus neutralising antibody titres pre and post booster vaccine, tested against D614G, delta, beta (and other VOCs should these become locally relevant) |
| | Evaluate cross-reactivity of T cell responses to ancestral and novel SARS-CoV-2 variants compared to baseline | T cell responses to D614G, delta, beta (and other VOCs should these become locally relevant) |
| **Exploratory** | Evaluate differences in clotting profiles with different boosters, age, and HIV status | Compare differences between haemoglobin, platelets and D-dimer at baseline and 2 weeks in each study arm and according to HIV status and age |

### 6. Methodology

#### 6.1. Trial design

This is a phase II randomised open label clinical trial in health care workers and other participants, age ≥18 years, who have previously received only one dose of the J&J Ad26.COV2.S vaccineand no other booster vaccines. PLWH and HIV-uninfected participants will be enrolled. We will aim to enrol at least 10% of participants ≥55 years and participants who may have known, well controlled comorbidities. Previous SARS-CoV-2 infection, prior to or after the J&J Ad26.COV2.S vaccine, will not result in exclusion but study results will be stratified according to evidence of previous infection. Participants will be recruited from 4 clinical trial sites in South Africa over a 4-month period. They will be recruited from 4 months after receiving the prime J&J Ad26.COV2.S vaccine.

Participants will be randomised 1:1:1:1 to group A, B, C and D. Group A will receive the Ad26.COV2.S prime plus full dose Ad26.COV2.S booster at a dose of 5x $10^{10}$ vp/ml (0.25 ml); Group B will receive the Ad26.COV2.S prime plus half dose Ad26.COV2.S booster at a dose of 2.6x $10^{10}$ vp/ml (0.13 ml); Group C will receive Ad26.COV2.S prime plus full dose BNT162b2 booster at a dose of 30 mcg (0.3 ml); and Group D will receive Ad26.COV2.S prime plus half dose BNT162b2 booster at a dose of 15 mcg (0.15 ml). Participants will be followed up for 6 months on study after randomisation.

#### 6.2. Study groups

Table 5: Study groups for the J&J Ad26.COV2.S/Pfizer BNT162b2 prime-boost study

| Group | Number | Time after J&J Ad26.COV2.S prime | Objectives | Vaccination | Vaccine Schedule |
|---|---|---|---|---|---|
| A<br>N= 75 | 50 HIV Uninfected<br>25 PLHIV* | ≥ 4 months | Immunogenicity and safety | J&J Ad26.COV2.S<br>5x $10^{10}$ vp/ ml | One dose<br>(0.25 ml) |
| B<br>N=75 | 50 HIV Uninfected<br>25 PLHIV* | ≥ 4 months | Immunogenicity and safety | J&J Ad26.COV2.S<br>2.6x $10^{10}$ vp/ ml<br>(dose for rounded off volume) | One dose<br>(0.13 ml – rounded off for ease of correct dosing) |
| C<br>N=75 | 50 HIV uninfected<br>25 PLHIV* | ≥ 4 months | Immunogenicity and safety | Pfizer BNT162b2 30 mcg IMI | One dose<br>(0.3 ml) |
| D<br>N=75 | 50 HIV uninfected<br>25 PLHIV* | ≥ 4 months | Immunogenicity and safety | Pfizer BNT162b2 15 mcg IMI | One dose<br>(0.15 ml) |

*At least a third of participants will be living with HIV, up to 150 participants living with HIV can be recruited across the study.

An interim analysis for immunogenicity will be done after the 3-month visit. If insufficient immunogenicity (defined as <75% of the geometric mean titre (GMT)) is observed between the full dose and half dose arms, or between J&J Ad26.COV2.S and Pfizer BNT162b2 arms, regardless of dose, the homologous or heterologous booster vaccine eliciting the most robust response will be offered at the 6-month visit. An interim analysis for immunogenicity will be performed after the 3-month visit. The highest GMT of neutralising antibodies, calculated per arm for each of the four arms, will be used as the benchmark for comparison to individual immunogenicity responses. The GMT of neutralising antibodies from the 3-month post-vaccination visit will be utilised for this purpose. If insufficient immunogenicity (defined as <75% of the geometric mean titre (GMT)) is observed between the full dose and half dose arms, or between J&J Ad26.COV2.S and Pfizer BNT162b2 arms, regardless of dose, the homologous or heterologous booster vaccine eliciting the most robust response will be offered at the 6-month visit. Any participant, in any study arm (including the arm that elicits the highest GMT), will be offered the booster that elicited the highest GMT, if their 3-month neutralising antibody level is <75% of the highest GMC. Participants will be contacted by the study staff and will be offered a booster dose if these criteria are met. The study Data and Safety Monitoring Board will review data and adjudicate on this process. Participants who receive a second boost at 6 months will receive a diary card to complete and a telephonic review on day 7 post vaccine. An interim visit will be arranged if previous criteria for an interim visit are met.

#### 6.3. Trial participants

Study participants will be ≥ 18 years, aiming to recruit at least 10% ≥ 55 years, and at least 1/3 PLHIV and 2/3 HIV-uninfected. Participants may enrol if they have no or well controlled comorbidities and have only had 1 dose of the J&J Ad26.COV2.S vaccine, without any further boosting. Participants will not be screened for previous SARS-CoV-2 infection prior to enrolment but will have nasopharyngeal PCR testing on the day of enrolment. Participants will also have nucleocapsid antibody testing at enrolment to identify those previously infected with SARS-CoV-2, but these results will not be available prior to vaccination and will be noted when analysing response results. Participants will be recruited ≥4 months after receiving the J&J Ad26.COV2.S prime.

#### 6.4. Potential risks for participants

Risks may be associated with phlebotomy, vaccine reactogenicity, anaphylaxis, excessive response to the booster vaccine, and disclosure of HIV status.

##### 6.4.1. Venipuncture

Bruising or swelling may occur as a result of venipuncture. Across the course of the study 10-70ml of blood will be taken per visit (SOE Table 3), in total less than 200 ml over 6 months which is an acceptable volume according to the Office for Human Research Protections (OHRP). he recommended maximum volume of blood taken in “healthy, nonpregnant adults who weigh at least 110 pounds” is that “the amounts drawn may not exceed 550 ml in an 8 week period and collection may not occur more frequently than 2 times per week”(49).

##### 6.4.2. Vaccine reactions

###### *6.4.2.1. Local reactions*

May include pain, tenderness, redness or swelling at the vaccination site.

###### *6.4.2.2. Systemic reactions*

May include, but not limited to fever, malaise, tiredness, headache, fatigue, myalgia, chills, cough and loss of taste and smell, and usually last 2-3 days post booster vaccination.

Rare thrombocytopenic thrombosis mediated by platelet activating antibodies against platelet factor have been described in a small number of people receiving the ChAdOx1 nCov-19 vaccine. This has resulted in presentations including cerebral venous thrombosis, splanchnic thrombosis and pulmonary embolism occurring 1-2 weeks post vaccination (50, 51). These findings raised concerns about potential similar risks in other adenovirus-based vaccines including the J&J Ad26.COV2.S vaccine. In the Sisonke IIIB study 5 health care workers, all who had risk factors, experienced thromboembolic events, a case rate of 1.7/100 000 (13). Two cases of thrombocytopenic thrombosis related to the J&J Ad26.COV2.S have been reported in the SISONKE Phase IIIB study.

With the Pfizer BNT162b2 vaccine, anaphylactic responses have also been reported worldwide as a rare event, with 175 cases of severe allergy with 21 cases (11.1/1 million) confirmed to be anaphylaxis meeting the Brighton collaboration case definition criteria (23). Myocarditis has been reported with the mRNA vaccines, at a frequency of 39–47 cases/million, more commonly in males younger than 30 years (22) with the majority of cases being self-limiting. Syncope post vaccination may also occur in rare cases.

###### 6.4.3. Response to booster

It is possible that participants who receive the booster may have an enhanced response to the vaccine resulting in excess systemic symptoms.

###### 6.4.4. Anaphylactic reactions

Anaphylactic reactions to the vaccines are possible but extremely rare. The clinical teams at each site are trained in advanced clinical life support (ACLS) and are able to manage and stabilise a participant who develops anaphylaxis, prior to transfer to hospital care for further observation and management.

###### 6.4.5. Loss of privacy or confidentiality

HIV testing will be done at study screening/entry visit. An HIV test will be conducted for all participants not known to be PLHIV, and have either previously tested HIV-negative or have an unknown HIV status. PLHIV will not be re-tested for HIV. In the situation where a person tests HIV positive, there is a small risk that HIV status may result in loss of confidentiality, stigma, or a lack of privacy. This will be avoided as far as possible by confidential management of all study source notes and documentation, use of a participant identifier and research staff following a strong code of principles regarding maintenance of privacy and confidentiality. PLHIV who are newly diagnosed will remain on study, be counselled about their HIV status, and be referred to HIV clinical and ART services. People living with HIV and those who are HIV-uninfected will have the same study visits conducted to minimise any potential loss of confidentiality.

##### 6.5. Known potential benefits

This study is evaluating four booster vaccine options, the J&J Ad26.COV2.S homologous boost at full dose (5x $10^{10}$ vp/ml, 0.25 ml) and half dose (2.6x $10^{10}$ vp/ml, 0.13 ml) and the Pfizer BNT162b2 at full dose (30mcg, 0.3 ml) and half dose (15mcg, 0.15 ml) ≥ 4 months after the J&J Ad26.COV2.S vaccine prime. Although this study is experimental, we expect that each of the booster doses will demonstrate a robust immune response and therefore there is a direct benefit to study participation.

Although the prime-boost strategy with J&J Ad26.COV2.S and Pfizer BNT162b2 has not yet been studied, data from a similar Adenovirus-vectored prime, the ChAdOx1 vaccine, and Pfizer BNT162b2 vaccine boost has been studied with promising results (41). Recent data exploring a quarter dose J&J Ad26.COV2.S booster at 6 months post prime demonstrated robust antibody responses at day 28 post boost, comparable to levels at day 29 post prime (15). The Pfizer BNT162b2 15mcg dose is also expected to result in a robust immune response when used as a booster, although no published data is available regarding this strategy. In the phase I Pfizer BNT162b2 study, a 20mcg dose of vaccine demonstrated a robust immune response compared to the full 30mcg dose response, suggesting that this strategy might be advantageous if similar responses are seen, particularly given existing constraints regarding vaccination supply and cost (19). An interim analysis for immunogenicity will be performed after the 3-month visit. The highest GMT of neutralising antibodies, calculated per arm for each of the four arms, will be used as the benchmark for comparison to individual immunogenicity responses. The GMT of neutralising antibodies from the 3-month post-vaccination visit will be utilised for this purpose. If insufficient immunogenicity (defined as <75% of the geometric mean titre (GMT)) is observed between the full dose and half dose arms, or between J&J Ad26.COV2.S and Pfizer BNT162b2 arms, regardless of dose, the homologous or heterologous booster vaccine eliciting the most robust response will be offered at the 6-month visit. Any participant, in any study arm (including the arm that elicits the highest GMT), will be offered the booster that elicited the highest GMT, if their 3-month neutralising antibody level is <75% of the highest GMC. Participants will be contacted by the study staff and will be offered a booster dose if these criteria are met. The study Data and Safety Monitoring Board will review data and adjudicate on this process. Final analysis will include the 6-month data.

#### 6.6. Recruitment and Withdrawal of trial participants

##### 6.6.1. Identification of trial participants

HCW's who participated in the SISONKE phase IIIB implementation study, as well as people vaccinated through the SA vaccination programme will be recruited through the following mechanisms:

- A bulk SMS containing brief details of the study will be sent to HCW on the SISONKE phase IIIB implementation study to inform them of this study and to invite them to contact the participating sites if interested.
- Advertisements through social media (Facebook), study site websites and pamphlets (paper and electronic).
- Word-of-mouth recruitment.
- Existing Sisonke sub-study cohort participants may also be approached to participate in the booster study.

Interested participants will contact the participating sites telephonically or by email and will be invited to site for an in-person visit where the participant will sign an informed consent form (ICF) prior to other screening procedures. Appendices 1, 2 and 3.

##### 6.6.2. Informed consent process

Each participant will be given the opportunity to read through the ICF. The participant will have the opportunity to ask any questions regarding the ICF and the study with the research team and will also be able to take the ICF home for further discussion with family or other support whilst considering participation.

Comprehension will be assessed and the ICF will be signed before any study procedures are conducted. ICFs will be offered in English, with strict adherence to GCP principles relating to the use of translators and witnesses for English-illiterate volunteers. For the purpose of this study, only English ICFs will be available.

The following principles will be emphasized during the ICF discussion:

- Participation in the study is entirely voluntary;
- Refusal to participate involves no penalty or loss of medical benefits;
- The participant has the right to withdraw participation at any time;
- The participant may ask questions at any time to increase understanding of the purpose of the study and the procedures involved;
- The study is investigating four dosing approaches to COVID-19 vaccine boosting of the J&J Ad26.COV2.S vaccine given through the SISONKE phase IIIB implementation study or the SA vaccination programme, namely the J&J Ad26.COV2.S vaccine (5x $10^{10}$ vp/ml)

or half dose (2.6x $10^{10}$ vp/ml) or the Pfizer BNT162b2 vaccine at 30mcg or 15mcg doses;

- Since a booster vaccine is being given to participants, there is an expected direct benefit to participating in the study, however the extent of this benefit is being evaluated;
- Participants will be asked to provide detailed medical and surgical history to investigator verbally and if possible, patient-held medical records (outpatient cards) will be reviewed;
- Documentation proving the identity of the participant will be required and confidentiality will be maintained;
- Participants will need to demonstrate proof of participation in SISONKE phase IIIB implementation study or SA vaccination programme, by showing a valid SISONKE or DoH vaccination card;
- A Biometric Co-Enrolment Prevention System or similar process will be used by all sites to prevent co-enrolment in other interventional studies.

##### 6.6.3. Inclusion and exclusion criteria

###### *6.6.3.1. Inclusion criteria*

All inclusion criteria for the study must be met for participation:

1. HCW's and others who received the J&J Ad26.COV2.S vaccine through the SISONKE phase IIIB implementation study or the SA vaccination programme .
2. Age ≥ 18 years.
3. HIV-uninfected or PLHIV medically stable on the day of enrolment.
4. Female participants of childbearing potential must have a negative urine BHCG test at screening/enrolment.
5. Willing and able to sign informed consent.
6. Able in the investigator's opinion to comply with study procedures.
7. Plan to stay within reasonable distance of the study site to attend study visits.
8. No comorbidities, or well controlled comorbidities, ≤ grade 1 severity at enrolment, including hypertension, type 1 or 2 diabetes, asthma or chronic pulmonary disease, tuberculosis in preceding 3 years, renal disease, cardiac conditions, or autoimmune conditions.

###### *6.6.3.2. Exclusion criteria*

1. Prior history of thrombotic events including previous deep vein thrombosis.
2. Receipt of any oral or other systemic steroid therapy within the preceding 28 days prior to enrolment.
3. Receipt of any other systemic immunosuppressive agent in the preceding 60 days prior to enrolment.
4. Receipt of any blood transfusion or any blood products within the preceding 6 months prior to enrolment.
5. Receipt of any vaccines in the 28 days prior to enrolment.
6. Cancer either not in remission, or not in remission in preceding 5 years prior to enrolment.
7. Any history of allergic disease or reactions likely to be exacerbated by any component of the vaccine.
8. Any history of hereditary angioedema or idiopathic angioedema.
9. Any history of anaphylaxis secondary to vaccination.
10. Current pregnancy at enrolment.

11. Receipt of any COVID-19 vaccine except for the J&J Ad26.COV2.S vaccine through the SISONKE phase IIIB implementation study or SA vaccination programme.
12. Symptoms of SARS-CoV-2 disease at the screening /enrolment visit will result in exclusion.
13. Participants not willing to be tested for HIV at entry, unless known to be PLHIV, will be excluded.
14. Enrolment in any other investigational studies for duration of the BaSiS study.

###### 6.6.4. Female participants of childbearing potential

Female participants of childbearing potential will be required to have a negative urine pregnancy test at screening/enrolment. Given that tens of thousands of pregnant women have been vaccinated with the J&J Ad26.COV2.S and Pfizer BNT162b2 vaccine globally, and that no safety concerns have been raised, contraception will not be required. National guidelines from Department of Health recommend vaccination for all pregnant women with either of the currently available vaccines. However, as there is insufficient data on the safety of booster doses in pregnant women, they will also be encouraged to defer pregnancy for at least 8 weeks post vaccination given limited data in early pregnancy.

##### 6.7. Trial procedures

All participants will attend study scheduled visits until 6 months post-randomisation and booster vaccination. HIV-uninfected participants and those living with HIV will have a combined screening and enrolment visit conducted on the same day. PLHIV will be eligible if they are medically stable with no acute illness or comorbidities that in the investigator's opinion will compromise their safety.

Study visit procedures for HIV-uninfected and PLHIV participants are tabulated in table 3. Study visits occur at screening and enrolment, 2 weeks, 3 months, and 6 months. Blood samples at each study visit will be taken, up to approximately 70 ml per visit, < 250 ml across the study. At the screening/enrolment visit SARS-CoV-2 nasopharyngeal swab testing will be conducted but results will not be required before enrolment and vaccination. If a participant tests positive, they will be evaluated in a subset of the study. Antibody testing done as part of the baseline study procedures will evaluate previous SARS-CoV-2 infection with nucleocapsid testing, however prior infection will not be an exclusion and stratification will be done as part of the analysis. Participants requiring an additional booster will be contacted to offer the booster, and will receive the booster at approximately 6 months (+ 2m) after enrolment.

###### 6.7.1. Observations

At each visit vital signs including temperature, heart rate, respiratory rate, oxygen saturation on room air and blood pressure will be measured. A medical history will be taken including questions regarding recent COVID-19 close contact, COVID-19 symptoms, positive SARS-CoV-2 antigen or nasopharyngeal polymerase chain reaction test and a targeted physical examination conducted if necessary.

###### 6.7.2. Spike binding antibody assays

Spike binding antibody assays will be performed at enrolment, 2 weeks, 3 months and 6 months. Assays will be performed as follows: 2mcg/ml of ancestral D614G spike protein will be used to coat 96-well, high-binding plates and incubated overnight at 4°C. The plates will be incubated in a blocking buffer consisting of 5% skimmed milk powder, 0.05% Tween 20, 1x PBS. Plasma samples will be diluted to 1:100 starting dilution in a blocking buffer and added to the plates. Secondary antibody will be diluted to 1:3000 in blocking buffer and added to the plates followed by TMB substrate (Thermofisher Scientific). Upon stopping the reaction with 1M $H_2SO_4$, absorbance will be measured at a 450nm wavelength. In all instances, mAbs CR3022 and BD23 will be used as positive controls and palivizumab was used as a negative control. All values will be normalized with the CR3022 mAb(52).

##### 6.7.3. Neutralisation Assays

###### *6.7.3.1. Pseudovirus neutralisation assays*

These will be performed on spike positive samples at enrolment, 2 weeks, 3 months, and 6 months. SARS-CoV-2 pseudotyped lentiviruses will be prepared by co-transfecting the HEK 293T cell line with either the SARS-CoV-2 ancestral variant spike (D614G), the Beta spike (L18F, D80A, D215G, K417N, E484K, N501Y, D614G, A701V, 242-244 del) or the Delta spike (T19R, R158G L452R, T478K, D614G, P681R, D950N, 156-157 del) plasmids in conjunction with a firefly luciferase encoding lentivirus backbone plasmid. As other VOC emerge these may be included in this panel. For the neutralisation assay, heat-inactivated plasma samples from vaccine recipients will be incubated with the SARS- CoV-2 pseudotyped virus for 1 hour at 37°C, 5% $CO_2$. Subsequently, $1x10^4$ HEK 293T cells engineered to over-express ACE-2 will be added and incubated at 37°C, 5% $CO_2$ for 72 hours upon which the luminescence of the luciferase gene was measured. CB6 will be used as a positive control.

###### *6.7.3.2. Live virus neutralisation assays*

These will be performed at enrolment, 2 weeks, 3 months and 6 months. Vero E6 cells will be plated in a 96-well plate at 30,000 cells per well 1 day before infection. Plasma will be separated from EDTA-anticoagulated blood by centrifugation at 500 rcf for 10 minutes and stored at −80°C. Aliquots of plasma samples will be heat-inactivated at 56°C for 30 minutes and clarified by centrifugation at 10,000 rcf for 5 minutes, after which the clear middle layer will be used for experiments. Inactivated plasma will be stored in single-use aliquots to prevent freeze–thaw cycles. For experiments, plasma will be serially diluted twofold from 1:100 to 1:3,200. As a positive control, the GenScript A02051 anti-spike monoclonal antibody will be added. Virus stocks will be used at approximately 100 focus-forming units per microwell and added to diluted plasma; antibody–virus mixtures will be incubated for 1 hour at 37°C, 5% $CO_2$. Cells will be infected with 100µl of the virus–antibody mixtures for 1 hour, to allow adsorption of virus. Subsequently, infection will be overlaid with 1.5% carboxymethylcellulose, Infection will be fixed with 4% PFA after 24 hours and imaged using an Elispot reader or automated microscope. SARS-CoV-2 variants will include sequenced outgrowths of ancestral SARS-CoV-2 with D614G only, beta, delta, and new variants to emerge.

##### 6.7.4. T cell assays

###### *6.7.4.1. Cell stimulation and flow cytometry staining*

Cryopreserved PBMC will be thawed, washed and rested in RPMI 1640 containing 10% heat-inactivated FCS for 4 hours prior to stimulation. PBMC will be seeded in a 96-well V-bottom plate at ~$2 \times 10^6$ PBMC per well and stimulated with SARS-CoV-2 spike peptide pool based on the full spike protein (Miltenyi Biotec), at 1µg/ml. A subset of samples will be tested for cross-reactivity of T cells to beta, delta, and new variants to emerge, using peptide pools based on these sequences. All stimulations will be performed in the presence of Brefeldin A (10µg/ml, Sigma-Aldrich, St Louis, MO, USA) and co-stimulatory antibodies against CD28 (clone 28.2) and

CD49d (clone L25) (1µg/ml each; BD Biosciences, San Jose, CA, USA). As a negative control, PBMC will be incubated with co-stimulatory antibodies, Brefeldin A and an equimolar amount of DMSO.

After 16 hours of stimulation, cells will be washed, stained with LIVE/DEAD™ Fixable VIVID Stain (Invitrogen, Carlsbad, CA, USA) and subsequently surface stained with the following antibodies: CD14 Pac Blue (TuK4, Invitrogen Thermofisher Scientific), CD19 Pac Blue (SJ25-C1, Invitrogen Thermofisher Scientific), CD4 PERCP-Cy5.5 (L200, BD Biosciences, San Jose, CA, USA), CD8 BV510 (RPA-8, Biolegend, San Diego, CA, USA), CD27 PE-Cy5 (1A4, Beckman Coulter), CD45RA BV570 (HI100, Biolegend, San Diego, CA, USA). Cells will be fixed and permeabilized using Cytofix/Cytoperm buffer (BD Biosciences) and stained with CD3 BV650 (OKT3), IFN-g Alexa700 (B27), TNF BV786 (Mab11) and IL-2 APC (MQ1-17H12) from Biolegend. Finally, cells will be washed and fixed in CellFIX (BD Biosciences). Samples will be acquired on a BD LSR-II flow cytometer and analyzed using FlowJo (v10, FlowJo LLC, Ashland, OR, USA). Cells will be gated on singlets, CD14- CD19-, live lymphocytes and memory cells (excluding naive CD27+ CD45RA+ population). Results will be expressed as the frequency of CD4+ or CD8+ T cells expressing IFN-g, TNF-a or IL-2. Parallel stimulation cultures will be set up in the absence of BFA and supernatant harvested for IFN-g ELISA.

##### 6.7.5. Study visits

###### *6.7.5.1. Screening and enrolment visit (Day 0)*

The screening and enrolment visit will be conducted together:

- Voluntary written informed consent will be obtained prior to commencement of study procedures. ICF may be conducted separately, and up to 6 weeks before the enrolment visit.
- Once ICF is obtained, vital signs will be measured, including blood pressure, pulse rate, oxygen saturation in room air, respiratory rate, oral temperature with a plastic sleeve over the thermometer or tympanic temperature with an infrared tympanic thermometer. Weight and height will be measured, and body mass index calculated.
- Targeted medical history for the preceding 5 years will be taken including any history of previous chronic diseases and previous anaphylaxis.
- Concomitant medication taken within the last 30 days will be recorded.
- COVID-19 history will be taken including history of previously confirmed SARS-CoV-2 infection by nasopharyngeal PCR or antigen testing, or a positive SARS-CoV-2 antibody test; close contact diagnosed with SARS-CoV-2 infection; symptoms since March 2020 that could be attributed to SARS-CoV-2 infection (including but not limited to fever or chills; cough; shortness of breath or breathing difficulties; fatigue; muscle or body aches; headache; loss of taste or smell newly developed; sore throat; congestion or runny nose; nausea or vomiting and diarrhoea).
- Targeted physical examination.
- Contraceptive history and urine pregnancy testing for all female participants of child-bearing potential.
- HIV Elisa test.
- Participants who are not known to be living with HIV and who test positive for HIV at this visit will receive counselling from a trained staff member. They will be referred to a practitioner or clinic of their choice for further management and for initiation of ART.

- Immunology blood tests as described in the SOE table 2.
- Immunogenicity bloods as described in SOE table 2.
- Randomisation 1:1:1:1 to group A, B, C or D.
- Vaccination.
- Observation for at least 15minutes post vaccination, including vital signs and assessment of local and systemic reactions
- Diary card instruction and issue, and thermometers will be issued with instructions regarding their use.

###### *6.7.5.2. Telephonic visit Day 7 (+ 3 days)*

A follow-up telephonic visit has been scheduled post-vaccination, to elicit if there are any adverse events (AE's) which occur within 7 days of booster vaccination, and if there are, to confirm if any AE's reported are grade 3 or higher, so that an interim visit can be scheduled. The telephonic visit will be conducted on day 7, with the day of vaccination counted as day 0. The window period for all telephonic visits post-vaccination is **+3 days**, with day 7 counted as the first day of the window period.

###### *6.7.5.3. Interim visits - only if indicated*

Interim visits will be conducted for the following reasons:

- Reactogenicity event greater or equal to grade 3 or any reactogenicity event ongoing at day 7.
- Solicited AEs beyond 7 days post vaccination, or unsolicited AEs throughout the study post vaccination if they are grade 3 or higher.
- Clotting abnormalities grade 2 or higher at baseline and week 2 or symptomatic between vaccination and the week 2 visit.

Participants will complete the diary card as instructed. A staff member will contact participants telephonically at day 7 to elicit if any grade 3 or higher AEs have occurred, or if AEs of any grade are noted to be ongoing at day 7. If present, the site will arrange an interim visit for the participant. Participants will be instructed to contact the site for any unsolicited AEs throughout the study if they are grade 3 or higher to arrange an interim visit.

For clotting abnormalities at baseline or week 2 follow-up, the site will contact the participant to enquire whether any symptoms are present, and arrange an interim follow up visit if grade 2 and symptomatic or grade 3 abnormalities. Repeat visits for ongoing abnormalities may be required.

For participants requiring an interim visit the following procedures will be performed:

- Collect and review diary card.
- Vitals.
- Medical and COVID-19 history (previously confirmed SARS-CoV-2 infection by NP PCR or antigen testing, or a positive SARS-CoV-2 antibody test; close contact diagnosed with SARS-CoV-2 infection; symptoms since March 2020 that could be attributed to SARS-CoV-2 infection (including but not limited to fever or chills; cough; shortness of breath breathing difficulties; fatigue; muscle or body aches; headache; loss of taste or smell newly developed; sore throat; congestion or runny nose; nausea or vomiting and diarrhoea).
- Concomitant medication review.

- Targeted physical examination if required.

###### 6.7.5.4. Week 2 visit (±3 days)

- Collect diary card for participants not requiring a day 7 visit.
- Vitals as previously explained.
- Medical and COVID-19 history (previously confirmed SARS-CoV-2 infection by NP PCR or antigen testing, or a positive SARS-CoV-2 antibody test); close contact diagnosed with SARS-CoV-2 infection; symptoms since March 2020 that could be attributed to SARS-CoV-2 infection (including but not limited to fever or chills; cough; shortness of breath breathing difficulties; fatigue; muscle or body aches; headache; loss of taste or smell newly developed; sore throat; congestion or runny nose; nausea or vomiting and diarrhoea).
- Diary AE review if not done at an interim visit.
- Concomitant medication review.
- Targeted physical examination if required.
- Immunology bloods.
- Immunogenicity bloods.

###### 6.7.5.5. Month 3 visit (±7 days)

- Diary card collection if AEs are ongoing at the 2-week visit.
- Vitals as previously explained.
- Medical and COVID-19 history as previously described.
- Concomitant medication review.
- Targeted physical examination if required.
- Immunology bloods.
- Immunogenicity bloods.

###### 6.7.5.6. Month 6 visit (±14 days), end of study visit

- Vitals as previously explained
- Medical and COVID-19 history as previously described.
- Concomitant medication review
- Targeted physical examination if required
- Immunology bloods
- Immunogenicity bloods
- People with an inadequate response to the booster will be offered a best-option booster (either full dose of their assigned vaccine or a different study vaccine) based on study findings
- Issue vaccine card for appropriate booster dose at exit visit.
- Telephone call at day 7 to collect diary care information and arrange additional visit to site, if necessary.

###### 6.7.5.7. Illness visits

An illness visit will be conducted for intercurrent SARS-CoV-2 symptoms or confirmed infection either as part of the enrolment visit, or after testing by a non-study provider. These visits will be conducted as soon as possible after the participant experiences symptoms or reports to have tested positive for SARS-CoV-2 infection. Management of SARS-CoV-19 disease will be the responsibility of the participant's attending doctor or clinic and the site will not take over management of these episodes. The participant will be seen once in an infection-controlled area within the clinic and the following procedures will be conducted:

- Vitals
- Medical and COVID-19 history as previously described
- Review of medical history
- Concomitant medication review
- AE review
- Targeted physical examination
- SARS-CoV-2 NP PCR test
- Only one illness visit is required for diagnostic purposes

*6.7.5.8 Repeat booster visit (6 months +2 months)*

Participants who meet study criteria for a repeat booster, will be contacted after the interim analysis is complete and will be offered the opportunity for an additional booster with the best booster as determined by the data and adjudication by the DSMB. This timing may vary by site due to staggered enrolment, but will ideally occur at 6 months post enrolment, or up to 8 months post enrolment. These participants will receive a diary card and will be contacted on day 7 post repeat booster as described in section 6.7.5.2. An interim visit will be arranged if required.

### 7. Investigational Product

The investigational product in the study includes the following:

- Johnson and Johnson adenovirus vector vaccine Ad26.COV2.S at full ($5.0 \times 10^{10}$) and half ($2.6 \times 10^{10}$) dose.
- Pfizer BNT162b2 mRNA vaccine by Pfizer BioNTech at full (30mcg) and half (15mcg) dose.

Participants will be randomised to one of four groups. Group A and B will receive the Johnson and Johnson adenovirus vector vaccine. Group C and D will receive the Pfizer mRNA vaccine. Each group will be assigned the investigational product at full, and half dose as detailed in the table below:

| | Group A | Group B | Group C | Group D |
|---|---|---|---|---|
| **Type of Vaccine** | Janssen Ad26.COV2.S | | Pfizer BNT162b2 | |
| **Formulation** | Adenovirus vector vaccine | | mRNA vaccine | |
| **Dosage** | $5.0 \times 10^{10}$ vp/ ml | $2.6 \times 10^{10}$ vp/ ml | 30mcg | 15mcg |
| **Diluent** | None required | | 0.9% Sodium chloride Injection USP. **No bacteriostatic 0.9% Sodium chloride should be used** | |
| **Final volume to be administered** | 0.25 ml | 0.13 ml | 0.30 ml | 0.15 ml |
| **Maximum doses per vial** | 2 | 4 | 6 | 12 |
| **Route of administration** | Intramuscular, deltoid muscle of non-dominant arm | | | |

#### 7.1. J&J Ad26.COV2.S adenovector vaccine

The J&J vaccine is composed of recombinant, replication-incompetent human adenovirus type 26 vector that, after entering human cells, expresses the SARS-CoV-2 spike S antigen without virus propagation. An immune response elicited to the S antigen protects against COVID-19. The vaccine will be supplied by SAMRC/Janssen as a frozen, multidose, preservative free suspension (0.50 ml).

#### 7.2. Pfizer BioNTech BNT162b2 modRNA vaccine

The Pfizer vaccine is formulated in lipid particles that enable delivery of the RNA into the host cells. The host cells allow expression of the SARS-CoV-2 S antigen. The vaccine elicits an immune response to the S antigen which protects against COVID-19. The vaccine will be supplied by the Department of Health as a frozen, multidose, preservative free suspension (0.45 ml). Each vial will require dilution with 0.9% sterile Sodium chloride (1.80 ml).

Refer to the investigational product (IP) manual for further clarification on study product.

#### 7.3. IP preparation, handling, storage and accountability

##### 7.3.1. J&J Ad26.COV2.S adenovector vaccine

The J&J vaccine is supplied as multi dose vials in cartons of 20. The cartons will arrive as frozen. Vials should be stored in a freezer between -15°C and -25°C. Vials must be kept frozen and protected from light. Vials must be kept in original cartons in the freezer in an upright position.

##### 7.3.2. Pfizer BioNTech BNT162b2 modRNA vaccine

The Pfizer BioNTech vaccine (COMIRNATY®) is supplied as multi dose vials in cartons of 25 or 195. The cartons will arrive in thermal containers with dry ice. Vials should be stored in an ultra-low freezer between -90°C to -60°C. Vials must be kept frozen and protected from light during storage.

The diluent (0.9% Sodium chloride injection USP) is provided separately and should be stored at room temperature (20-25°C). The provided diluent will be supplied as either 10 ml or 2 ml single use vials.

For both vaccines, the site pharmacist or designated study personnel will confirm if the study product has arrived at the appropriate temperature range as stipulated in the IP manual. The study product should be stored on site in a secure location and at the correct temperature according to manufacturers' recommendations, with limited access to unauthorized persons and at controlled temperatures as indicated on the clinical labels. Any temperature excursions should be reported as described in the IP manual.

Study vaccines will be prepared by appropriately qualified and delegated study personnel, and in accordance with the IP manual. Site pharmacists will be responsible for all study product accountability. All product received and dispensed must be accounted for using site-specific SOPs and accountability logs.

##### 7.3.3. IP destruction

The process of destruction of any unused study product and documentation of destruction is detailed in the IP manual and should be followed accordingly.

### 8. Assessment of safety

Safety will be assessed according to frequency, severity, and type of adverse event and severe adverse event. Local and systemic solicited and unsolicited reactogenicity adverse events will be collected following booster vaccine on day 0 until 28 days after vaccination.

The following solicited reactogenicity AEs will be collected on the dairy:

- Pain
- Tenderness
- Erythema/Redness
- Induration/Swelling
- Fever > 38.0 degrees Celsius
- Nausea/Vomiting
- Diarrhoea
- Headache
- Fatigue
- Myalgia
- Chills
- Cough
- Loss of taste and/or smell

SAEs and AESI will be collected from enrolment until study end, as well as any AEs resulting in withdrawal of the participant from the study.

#### 8.1. Definitions

##### 8.1.1. Adverse Event (AE)

Any untoward medical occurrence that occurs during the time of administration of a study product, including events related and not related to the study product.

##### 8.1.2. Adverse reaction (AR)

This is a reaction to the study product assessed to be related (definitely, probably or possibly) to the study product as evaluated by the investigator.

##### 8.1.3. Serious Adverse event (SAE)

This is an adverse event that results in one of the following outcomes:

- Death
- Life-threatening if no intervention
- Hospitalisation or prolonged hospitalisation
- Results in permanent or significant disability
- Congenital anomaly

##### 8.1.4. Important medically attended event

This is an adverse event that does not meet criteria for an SAE, SUSAR or AESI but is a medical visit for a reason other than routine study visit or vaccination and requires a hospital, emergency room or unscheduled visit to medical personnel (including the site) for any reason.

##### 8.1.5. Suspected Unexpected Severe Adverse Reaction (SUSAR)

A suspected unexpected serious adverse response to the study product, not included in the study product information brochure but meeting the criteria as defined in "Serious adverse event". Since no SAEs are expected as a result of booster vaccination in this study, all SAEs related to vaccine will be reported as SUSARs.

##### 8.1.6. Adverse Event of Special Interest (AESI)

AESIs will include the following AEs or SAEs:

- Moderate or severe SARS-CoV-2 infection resulting in hospitalisation, death or where residual complications occur such as oxygen dependency.
- Myocarditis, stroke or any other neurological complications of SARS-CoV-2 infection.

#### 8.2. Grading of events

The Guidance for Industry: Toxicity Grading Scale for Healthy Adult and Adolescent Volunteers Enrolled in Preventive Vaccine Clinical Trials (53)will be the primary grading table in keeping with the previous Pfizer COVID vaccine grading standards.

##### 8.2.1. Reactogenicity

A diary card will be issued to each participant on the day of randomisation and vaccination. The participant will record local and systemic side effects to the vaccine on the diary card.

###### *8.2.1.1. Local reactions*

Expected reactions include pain, tenderness, erythema/redness and induration/swelling. These local reactions will be graded according to FDA guidance below (53):

| **Local Reaction to Injectable Product** | **Mild (Grade 1)** | **Moderate(Grade 2)** | **Severe (Grade 3)** | **Potentially Life Threatening (Grade 4)** |
|---|---|---|---|---|
| Pain | Does not interfere with activity | Repeated use of non-narcotic pain reliever > 24 hours or interferes with activity | Any use of narcotic pain reliever or prevents daily activity | Emergency room (ER) visit or hospitalization |
| Tenderness | Mild discomfort to touch | Discomfort with movement | Significant discomfort at rest | ER visit or hospitalization |
| Erythema/Redness * | 2.5 – 5 cm | 5.1 – 10 cm | > 10 cm | Necrosis or exfoliative dermatitis |
| Induration/Swelling ** | 2.5 – 5 cm and does not interfere with activity | 5.1 – 10 cm or interferes with activity | > 10 cm or prevents daily activity | Necrosis |

A simplified version of these criteria will be included in the diary card.

###### *8.2.1.2. Systemic reactions*

Systemic reactions including fevers above 38.0°C, nausea, vomiting, diarrhoea, headache, fatigue myalgia, chills, cough, loss of taste or smell will be recorded on the diary card.

Anaphylaxis will be graded according to criteria included in Appendix 1.

Systemic reactions will be graded as follows (FDA)

| Systemic (General) | Mild (Grade 1) | Moderate(Grade 2) | Severe (Grade 3) | Potentially Life Threatening (Grade 4) |
|---|---|---|---|---|
| Nausea/vomiting | No interference with activity or 1 – 2 episodes/24 hours | Some interference with activity or > 2 episodes/24 hours | Prevents daily activity, requires outpatient IV hydration | ER visit or hospitalization for hypotensive shock |
| Diarrhea | 2 – 3 loose stools or < 400 gms/24 hours | 4 – 5 stools or 400 – 800 gms/24 hours | 6 or more watery stools or > 800gms/24 hours or requires outpatient IV hydration | ER visit or hospitalization |
| Headache | No interference with activity | Repeated use of non-narcotic pain reliever > 24 hours or some interference with activity | Significant; any use of narcotic pain reliever or prevents daily activity | ER visit or hospitalization |
| Fatigue | No interference with activity | Some interference with activity | Significant; prevents daily activity | ER visit or hospitalization |
| Myalgia | No interference with activity | Some interference with activity | Significant; prevents daily activity | ER visit or hospitalization |

| Systemic Illnes | Mild (Grade 1) | (Moderate(Grade 2) | Severe (Grade 3) | Potentially Life Threatening (Grade 4) |
|---|---|---|---|---|
| Illness or clinical adverse event (as defined according to applicable regulations) | No interference with activity | Some interference with activity not requiring medical intervention | Prevents daily activity and requires medical intervention | ER visit or hospitalization |

Expected systemic events will also be recorded in the diary card.

D-Dimer grading will be completed according to the FDA “Systemic Illness” table above. If D-Dimer levels are < 1.0 ug/ml they will not be considered significant or require further evaluation, unless any symptoms are present (personal communication Prof Barry Jacobson). An SOP is available for more detailed description of the follow-up process.

##### 8.2.2. Vital signs

Vital signs will be graded according to the FDA grading tables below.

| Vital Signs * | Mild (Grade 1) | Moderate(Grade 2) | Severe (Grade 3) | Potentially Life Threatening (Grade 4) |
|---|---|---|---|---|
| Fever (°C) **<br>(°F) ** | 38.0 – 38.4<br>100.4 – 101.1 | 38.5 – 38.9<br>101.2 – 102.0 | 39.0 – 40<br>102.1 – 104 | > 40<br>> 104 |
| Tachycardia - beats per minute | 101 – 115 | 116 – 130 | > 130 | ER visit or hospitalization for arrhythmia |
| Bradycardia - beats per minute*** | 50 – 54 | 45 – 49 | < 45 | ER visit or hospitalization for arrhythmia |
| Hypertension (systolic) - mm Hg | 141 – 150 | 151 – 155 | > 155 | ER visit or hospitalization for malignant hypertension |
| Hypertension (diastolic) - mm Hg | 91 – 95 | 96 – 100 | > 100 | ER visit or hospitalization for malignant hypertension |
| Hypotension (systolic) – mm Hg | 85 – 89 | 80 – 84 | < 80 | ER visit or hospitalization for hypotensive shock |
| Respiratory Rate – breaths per minute | 17 – 20 | 21 – 25 | > 25 | Intubation |

* Subject should be at rest for all vital sign measurements.
** Oral temperature; no recent hot or cold beverages or smoking.
*** When resting heart rate is between 60 – 100 beats per minute. Use clinical judgement when characterizing bradycardia among some healthy subject populations, for example, conditioned athletes.

###### 8.2.3. Causality

The site investigator will assess the relationship between the study vaccine and the AE and provide an assessment of relatedness. It must be determined whether a reasonable possibility exists that the vaccine caused or contributed to a SAE.

This is based on clinical judgement and factors include:

- Whether a temporal relationship existed between the event and receipt of the vaccine.
- Whether there is a plausible biological mechanism for the vaccine to have caused the AE.
- If there is any other possible cause for the AE.
- Whether similar previous reports of AEs with the vaccine have been recorded and reported.

The AE may be assessed to be *related* (definite, probable, possible) if it is possible that the vaccine caused the AE and *not related* (not related or not likely) if there is no reasonable possibility that the vaccine caused the AE.

If an investigator assesses the AE as “not related”, an alternative cause, diagnosis, or explanation must be provided. Causality must be re-assessed when new information is available and may require review and update of the AE if applicable.

##### 8.3. Reporting of AEs

All AEs which occur within 7 days of booster vaccination should be reported on the paper-based diary card. Interim visits will be scheduled if any grade 3 or higher AE is recorded by the participant. All participants will be contacted telephonically on day 7 to ensure that all such events are recorded, and interim visits scheduled as necessary.

SAEs will be reported to the study team within 24 hours of site notification, as well as SUSARS, ADRs and AESI.

SAEs assessed to be related to study product or the trial or resulting in hospitalisation or death of a participant will be reported to the relevant ethics committee(s) within stipulated timelines, and regulatory authority (SAHPRA) according to the following guidance(54):

- Deaths or life threatening, related and unexpected reactions to be reported within 7 days (preliminary)
- New information impacting on risk-benefit profile of product or conduct of trial must be reported within 3 days.
- Other important safety info within 15 days.

### 9. Data Management and Analysis

#### 9.1. Randomisation Procedure

- All participants, who enrol in the trial will be individually randomised to one of four groups:
  Group A: full dose J&J Ad26.COV2.S (5x $10^{10}$ vp/ml, 0.25 ml)
  Group B: half dose J&J Ad26.COV2.S (2.6x $10^{10}$ vp/ ml, 0.13 ml)
  Group C: full dose BNT162b2 vaccine (30mcg)
  Group D: half dose (15mcg) Pfizer BNT162b2 vaccine 300 participants will be included across 4 arms A, B, C and D, 75 participants per arm (50 HIV-uninfected and 25 PLHIV).
- We will aim to include at least 10% of participants ≥ 55 years of age.
- Randomisation will not be blinded; open label vaccination will be followed.
- The GraphPad Software online randomisation or similar package will be used to perform stratified randomisation. Per site, randomisation will be performed separately within each stratum of HIV status to achieve treatment assignment balance. Therefore, at each site, 25 HIV-positive and 50 HIV-uninfected participants will be randomly assigned to Group A, B, C or D; this will result in at least 100 HIV-positive participants (up to 150 PLHIV) and 200 HIV-uninfected participants across the study.
- Participants in each stratum will be allocated on a 1:1:1:1 ratio in masked block sizes, ensuring balance between the groups. The study statistician or epidemiologist will create the randomisation allocation lists for each group prior to study start to ensure that the correct number of participants are randomised to each group at each site.
- The randomisation allocation list will be uploaded to REDCap for electronic implementation. Once the study site and HIV status of the participant is entered into the REDCap Demographic Form, REDCap will apply the random allocation table and assign the participant to a study arm. This study arm will be shown on the REDCap Demographic Form. Once a PID has been allocated to a study arm, the study site and HIV status fields will be locked for future editing.
- Apart from the study Epidemiologist, who creates the randomisation allocation list, no other study staff will have access to this allocation list, thus substantially minimising the chances of being able to determine the allocation prior to enrolment.

#### 9.2. Sample size

We calculate the sample size necessary to detect an increase in antibody mediated neutralisation when an increase is present following booster vaccination with Pfizer BNT162b2 vaccine 15mcg IMI (half-dose) with 90% probability. Effect size for the calculation is fold-increase in neutralisation post-boost. Fold-change is assumed to be at least as large as observed with a homologous boost with a fractional dose of Ad26.COV2.S, which is 6 to 7.7-fold (15). Non-inferiority of a heterologous boost with a fractional Pfizer BNT162b2 dose is likely, due to the higher neutralisation response to this vaccine relative to Ad26.COV2.S(16).

Since this is a Phase II study with a fixed sample size of n=75 per prime/boost combination, the following effect sizes are illustrative for the study. For the primary outcome, the change in neutralising antibodies between baseline and 2-weeks post boost, the study, with n=75 participants per arm, will have 90% power to demonstrate a 4-fold increase in response at a 2.5% significance level for a paired t-test using a superiority margin of 0.  For 2-fold increase superiority margin the study, with n=75 participants per arm, will have 80% power for a 6-fold increase in neutralising antibodies at 2 weeks post boost for a paired t-test at a 2.5% significance level. The baseline sample size information is based on South African laboratory data of 84 participants of which 51 were vaccinated and infected with Covid and 33 vaccinated uninfected with Covid. For the PRNT50 assay the log10(GMT) of 1.89 with standard deviation of 1.246 was obtained. The correlation between baseline and 2-weeks post boost values was taken as 0.2 resulting in standard deviation estimate of the difference on the log10 scale of 1.474.

#### 9.3. Database development

A web-based database will be developed using REDCap (Research Electronic Data Capture), a secure, web-based application designed to support data capture for research studies (55). REDCap is a 'closed' system, which cannot be accessed without an individual username/password. There is SSL (Secure Sockets Layer) encryption, firewalls and separate application, and database servers, with connections between the two strictly regulated. Data is backed up on a separate physical machine, as well as running frequent offsite backups. Data collected at facilities will not be stored on local devices (laptops/tablets). All data are immediately uploaded to the REDCap server via a secure internet connection provided by the respective study sites – public or facility-based wi-fi or other connections will not be used.

REDCap has a fully traceable audit trail. High-level users may have access to view a log of all occurrences of data exports, design changes, record creation, updating & deletion, user creation, record locking, and page views. As such, any activity on the database can be traced back to an individual user.

All REDCap database users will be allocated specific roles that allow for defined access levels. For example: a data capturer will be able to create and edit participant data but will not be able to delete any records, export any data or adjust user rights. A data manager will be granted higher level access to allow him/her to create users, edit fields, delete records and perform data exports under the supervision of the epidemiologist. Study PIs will have full access to all modules of the database. These roles will be defined by the epidemiologist and study data manager in consultation with the study PIs.

All REDCap users are trained in GCP, Human Subjects Protection, and REDCap security. No staff member is allowed to share access credentials as they will then become liable for any activity logged against their user profile.

#### 9.4. Data Entry and Management

##### 9.4.1. Study-related Forms

Electronic forms will be developed directly on the REDCap database including the following:

- Demographic information form
- Case report forms (CRF) for each study visit (screening and enrolment, week 2, month 3, month 6, interim and illness visit
- Medical history log
- AE log

- Concomitant medication logs
- Diary card capture
- Laboratory Results
- Study disposition form

  Source notes will be created for each CRF as above.

##### 9.4.2. Data Entry and Processing

- A single unique participant identifying number will be allocated to each participant once the ICF has been signed.
- This unique identifier will be used for all source documentation, all laboratory tests and all data entry.
- The participant identifying number will be entered on the Screening and Enrolment Log. When a data capturer creates a record for the participant on the REDCap database, they will enter that identifying number into the database.
- Linkage of the unique participant identifier to the participant file number and facility will be recorded on the Screening and Enrolment Log. During data capture, research assistants, the data manager, and Wits RHI sub-investigators will have access to the data in order to capture data and to maintain data quality.
- The database will be updated with information from Source documentation which will contain no personally identifying information. All these documents are located in the participants file at the specific research site, which is stored under restricted access to study personnel. All information entered in the database, will be entered under the unique participant identifying number.
- All data will be entered directly onto the REDCap database.
- The database will:
  - Only accept variables within reasonable pre-defined ranges (e.g., CD4 0-2,000/µL).
  - Detect and prevent entry of inconsistencies (e.g., date of birth after date of HIV clinic enrolment).
  - Detect missing data fields and prevent omission of crucial data (e.g., missing date of ART initiation).
- All data will be stored on the online REDCap database and backed up on the Wits RHI server periodically; both of which can be accessed only using password-protected logins.
- Only the data manager and the study investigators will have access to accumulating data across all sites.

##### 9.4.3. Data Security

- Data security systems will comply with the requirements of the POPIA Act.
- All sites will ensure that antivirus software safeguards all devices and systems from viruses and other malware.
- All handheld devices will have all information wiped from memory and returned to factory settings at the end of the data collection process.
- Inoperable devices will have all information wiped from memory and returned to factory settings (if possible).
- Using REDCap databases further ensures that no participant information is stored on local devices.

- Data exported from REDCap for analysis purposes will be de-identified. This data will be stored on Wits RHI's OneDrive/SharePoint server that includes encryption linked to Authorised Directory user authentication, i.e., a user will not be able to access another user's OneDrive unless login details are shared. Sharing of login details is not permitted under Wits RHI's IT policy.

##### 9.4.4. Data Quality Control

On-site quality control (QC) will be performed by delegated staff according to the site SOP to ensure that the study is proceeding according to study protocols and Good Clinical Practice (GCP) requirements, the personnel are fulfilling their job descriptions and responsibilities, the data collected is accurate and complete, and confidentiality of participants is observed.

The respective site Data Manager/s will be responsible for ensuring that data quality is maintained and will review collected data on a weekly basis to ensure that all fields are collected appropriately in the correct format.

Data quality attributes that will be checked:

- Within range values
- Missing data
- Data formats

When discrepancies are noted, queries will be raised with the clinical staff (clinicians, study coordinators and nurses) and corrections will be completed on source documents and on the REDCap database. All errors will be corrected in real time in the source as they are discovered, and on the data base within 72 hours of source correction. A log will be kept of all changes and corrections.

##### 9.4.5. Data Transfer

- Tablet/Laptop devices will be accessed using password-protected logins and will communicate with REDCap using SSL (Secure Sockets Layer), a security technology for establishing an encrypted link between a web server and a browser. All data is immediately uploaded onto the REDCap server. Therefore, the risk of loss of data is almost non-existent in the case of a tablet being lost or stolen.
- All data will be stored on REDCap database and backed up on a Wits RHI cloud-based server, both of which can only be accessed with password-protected logins.
- Study statisticians will analyse the data and will not receive any individually identifiable data. Study statisticians will only have access to de-identified and de-linked data.
- Data will be de-identified by removing all personally identifying information from the database. If required, all dates will also be perturbed (date shifted) to further preserve the confidentiality of records.
- De-identified electronic data may need to be transferred from the central REDCap database to investigators or statisticians and vice versa. Data will never be transferred via email. Each investigator will be assigned a secure username and password with appropriate data management privileges to access data via the REDCap online platform.
- All final study databases will not contain any individually identifiable data.

- For any secondary analyses, permission to conduct the analysis must be sought from the Principal Investigators as well as regulatory authorities. No personally identifying information will be shared for secondary analyses – only de-identified, de-linked data.

#### 9.5. Data Analysis

This study is designed to test the primary hypothesis that the study prime/boost combination vaccination will result in a minimum 6-fold increase in neutralisation at 2 weeks and will be evaluated at a 2.5% one-sided significance level using a statistical test for paired data. An interim analysis for immunogenicity will be done after the 3-month visits. The highest GMT of neutralising antibodies, calculated per arm for each of the four arms, will be used as the benchmark for comparison to individual immunogenicity responses. Insufficient immunogenicity will be defined as <75% of the geometric mean titre (GMT)). The GMT of neutralising antibodies from the 3-month post-vaccination visit will be utilised to determine sufficient immunogenicity. A detailed statistical analysis plan (SAP) will be developed prior to any data analysis being conducted. Interim analysis will be performed at the 2-week post-boost timepoint and will inform sample size required for the following timepoints. Because of the time-sensitive nature of the results, conclusions from the interim analysis may be published before study completion.

The SAP will be agreed upon and signed off by all protocol co-chairs. All available data will be used in the analysis. Briefly, the data analysis will include the immunogenicity, reactogenicity and safety analyses described below.

##### 9.5.1. Immunogenicity analyses

For beta and delta variants of concern as well as other emerging variants of concern, we will compute the following:

- Geometric Mean Concentration (GMC) of SARS-CoV-2 serum neutralising antibody levels (using PNA and LVNA assays) and T-cell responses, among Group A participants, at baseline compared to 2-weeks, 3 months and 6-months post J&J Ad26.COV2.S $5x10^{10}$ vp/ml booster vaccination, overall, stratified by PLHIV and HIV-uninfected participants and stratified by participants ≥ 55 years and < 55 years.
- GMC of SARS-CoV-2 serum neutralising antibody levels (using PNA and LVNA assays) and T-cell responses, among Group B participants, at baseline compared to 2-weeks, 3-months and 6-months post J&J Ad26.COV2.S $2.5x10^{10}$ vp/ml booster vaccination, overall, stratified by PLHIV and HIV-uninfected participants and stratified by participants ≥ 55 years and < 55 years.
- GMC of SARS-CoV-2 serum neutralising antibody levels (using PNA and LVNA assays) and T-cell responses, among Group C participants, at baseline compared to 2-weeks, 3 months and 6-months post Pfizer BNT162b2 30mcg booster vaccination, overall, stratified by PLHIV and HIV-uninfected participants and stratified by participants ≥ 55 years and < 55 years.
- GMC of SARS-CoV-2 serum neutralising antibody levels (using PNA and LVNA assays) and T-cell responses, among Group D participants, at baseline compared to 2-weeks, 3-months and 6-months post Pfizer BNT162b2 15mcg booster vaccination, stratified by PLHIV and HIV-uninfected participants and stratified by participants ≥ 55 years and < 55 years.
- Comparison of GMC of SARS-CoV-2 serum neutralising antibody levels (using PNA and LVNA assays) and T-cell responses, at baseline, 2-weeks, 3-months and 6-months, between Groups A, B, C and D participants.
- Comparison of GMC of SARS-CoV-2 serum neutralising antibody levels (using PNA and LVNA assays) and T-cell responses, among Group C and D participants, at baseline compared to 2-weeks, 3-months and 6-months post heterologous Pfizer BNT162b2 booster vaccinations, stratified by

duration of time between prime and boost (<6 months vs ≥ 6 months) to assess whether duration of time between prime and heterologous boost impacts immunogenicity.

Non-normally distributed data will be log-transformed prior to analysis. The GMC and associated 95% confidence intervals will be computed for each group at all time points noted above, by computing the anti-log of the mean difference of the log-transformed data. Geometric mean fold rises from baseline and corresponding 95% confidence intervals will be computed. Baseline levels will be established at the enrolment visit. Graphical representations of immunologic parameters will also be produced, as appropriate.

##### 9.5.2. Safety and Reactogenicity analyses

Counts and percentages of every local and systemic adverse reactions reported through diary cards, monitor ARs, SAEs, SUSARs or AESIs, will be reported per group and stratified by HIV status

#### 11. Appendices

1. Brighton Collaboration Case Definition of Anaphylaxis for use in *SPSU* study
2. Toxicity Grading Scale for Healthy Adult and Adolescent Volunteers Enrolled in Preventive Vaccine Clinical Trials, September 2007
